# Genome-wide association studies of lifetime and frequency cannabis use in 131,895 individuals

**DOI:** 10.1101/2024.06.14.24308946

**Authors:** Hayley H A Thorpe, Pierre Fontanillas, John J Meredith, Mariela V Jennings, Renata B Cupertino, Shreya Pakala, 23andMe Research Team, Sarah L Elson, Jibran Y Khokhar, Lea K Davis, Emma C Johnson, Abraham A Palmer, Sandra Sanchez-Roige

## Abstract

Cannabis is one of the most widely used drugs globally. Decriminalization of cannabis is further increasing cannabis consumption. We performed genome-wide association studies (**GWASs**) of lifetime (*N=*131,895) and frequency (*N=*73,374) of cannabis use. Lifetime cannabis use GWAS identified two loci, one near *CADM2* (rs11922956, *p*=2.40E-11) and another near *GRM3* (rs12673181, *p*=6.90E-09). Frequency of use GWAS identified one locus near *CADM2* (rs4856591, *p*=8.10E-09; *r*^2^=0.76 with rs11922956). Both traits were heritable and genetically correlated with previous GWASs of lifetime use and cannabis use disorder (**CUD**), as well as other substance use and cognitive traits. Polygenic scores (**PGSs**) for lifetime and frequency of cannabis use associated cannabis use phenotypes in *AllofUs* participants. Phenome-wide association study of lifetime cannabis use PGS in a hospital cohort replicated associations with substance use and mood disorders, and uncovered associations with celiac and infectious diseases. This work demonstrates the value of GWASs of CUD transition risk factors.

## Introduction

Approximately 209 million people globally reported using cannabis in 2020^1^. The number of people who use cannabis regularly is expected to increase as cannabis is decriminalized in many jurisdictions^2–4^. While people report using cannabis for medicinal purposes^5^, there is increasing evidence that cannabis use has short- and long-term adverse consequences across psychiatric, cognitive, and physical health^6–14^. Up to 27% of those who use cannabis in their lifetime are estimated to develop cannabis use disorder (**CUD**)^15^, in which cannabis use becomes problematic to an individual’s intra- and interpersonal wellbeing^16^. However, it is currently unclear what factors contribute most to the development of CUD, and thus of clinical interest to identify what makes an individual vulnerable to cannabis use and its negative effects.

Problematic cannabis use is estimated to be 51-78% heritable based on twin studies^17–19^ and recent genome-wide association studies (**GWASs**) have implicated hundreds of loci associated with CUD^20–23^. While CUD GWASs are of paramount importance, they come with three major caveats. First, these studies only examine one extreme of the addiction spectrum and neglect other substance-related behaviors and stages between substance initiation to substance use disorder (**SUD**) diagnosis (e.g., recreational use, escalating intake, dependence)^24^. These pre-addiction phenotypes^25^ are thought to dictate an individual’s progression to SUD^26–32^ and are heritable^17,26,31,33^. However, aside from GWASs of lifetime cannabis use (having *ever* versus *never* used cannabis in one’s lifetime) using data from the International Cannabis Consortium (**ICC**) and other sources^34,35^, which represents the opposing end of the addiction spectrum to CUD, the genetics of other pre-addiction cannabis traits are understudied^36,37^. Second, only a portion of those engaging in frequent cannabis use seek treatment or have a CUD diagnosis^38,39^. It is therefore unlikely that CUD GWASs and downstream analyses fully characterize the genetics of regular, potentially harmful cannabis use and its relationships with physical and mental health. Third, curating case/control SUD GWASs are costly and laborious because they require individual psychological assessments for both cases and controls. Pre-addiction phenotypes can be rapidly and inexpensively collected in large population-based cohorts via self-report questionnaires^40^.

We collected data from 23andMe, Inc. research participants by asking if they had ever used cannabis (*N*=131,895). Those who responded yes were asked a follow-up question about the number of days they used cannabis in their heaviest use period (*N*=73,374) as a measure of cannabis use frequency. We performed GWASs of lifetime and frequency of cannabis use, followed by a battery of secondary analyses to compare biological, genetic, and phenotypic associations. Because the frequency of cannabis use phenotype better distinguished between light and heavy use, we hypothesized that the genetics of frequency of cannabis use would more closely resemble CUD compared to lifetime cannabis use genetics.

## Results

### GWASs of Lifetime Cannabis Use and Frequency of Cannabis Use Uncover Associations with CADM2 and GRM3

Participant demographics are described in **Supplementary Table 1**. The cohort was 65.2% female with a mean age of 52.8±0.04 years old. Participant responses to surveys about lifetime and frequency of cannabis use are available in **Supplementary Table 2** and **Supplementary Fig. 1**.

For single nucleotide polymorphisms (**SNPs**) quality control, see **Supplementary Table 3**. Genomic control inflation factors for lifetime cannabis use (λ=1.08) and frequency of cannabis use (λ=1.03) suggested no substantial inflation due to population stratification for either GWAS. SNP-based heritability (***h2_SNP_***) was 12.88%±0.97 for lifetime cannabis use, greater than the *h2_SNP_* for lifetime cannabis use from the ICC (*h2_SNP_* =6.63%±0.43)^34^. *h2_SNP_* for frequency of cannabis use was 4.12%±0.72 (**Supplementary Table 4**).

We identified two genome-wide significant (*p*<5.00E-08) loci for lifetime cannabis use on chromosomes 3 and 7 (**Fig 1A**, **Supplementary Fig. 2-3**, **Supplementary Table 5**). The most significant association was with rs11922956 (*p*=2.40E-11, chr3p12.1) located upstream the Cell adhesion molecule 2 gene (*CADM2*), replicating findings from previous lifetime use^34^ and CUD^22,23^ GWASs. *CADM2* encodes a glycoprotein primarily expressed in the brain with functions in cell-cell adhesion, synaptic formation, excitatory neurotransmission, and energy homeostasis ^41,42^. We also found a novel association between lifetime cannabis use and rs12673181 (*p*=6.90E-09, chr7q21.11), which is a SNP upstream of Metabotropic glutamate receptor 3 gene (*GRM3*) encoding mGlu_3_. mGlu_3_ is an inhibitory group II receptor affecting a range of intracellular signaling cascades and cellular processes like glutamate neurotransmission and long-term plasticity ^43^.

**Figure 1.**
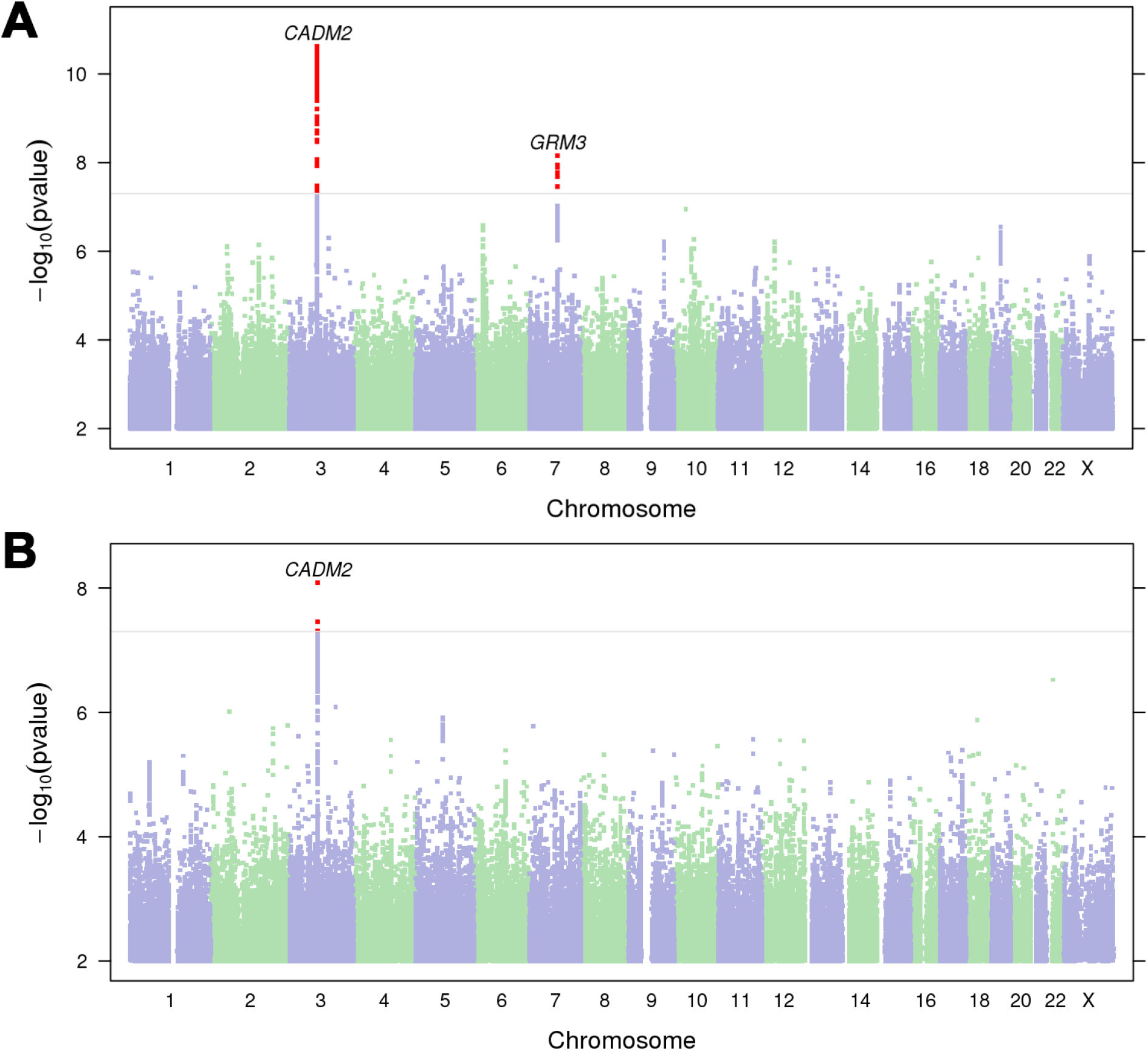
Manhattan plots of **A)** lifetime cannabis use (*N*=131,895) and **B)** frequency of cannabis use (*N*=73,374). The horizontal line represents the significance threshold (*p=*5.00E-08). Nearest protein-coding genes (<1Mb) to significant loci (red dots) are labelled. For quantile-quantile plots and locus zoom plots, see **Supplementary** Fig. 2-4.

Frequency of cannabis use GWAS identified one significant association with rs4856591 (*p*=8.10E-09, chr3p12.1; **Fig1B**, **Supplementary Fig. 2, 5**), which is near to *CADM2* and is in linkage disequilibrium (**LD**) with rs11922956 (*r*^2^=0.76, *p*<1.00E-04).

### Secondary Analysis Identifies 40 Lifetime and 4 Frequency of Cannabis Use Genes

Mapping SNPs to genes via gene-based (i.e., **MAGMA**, **H-MAGMA**) and transcriptome-wide association study (**TWAS**; i.e., S-PrediXcan) analyses identified 40 unique genes associated with lifetime cannabis use (**Supplementary Tables 6-8**), and 4 unique genes associated with frequency of cannabis use (**Supplementary Tables 9**). None of the 4 genes associated with frequency of cannabis use (i.e., *MMS22L*, *DSCC1*, *CPSF7*, *RP11-51J9.6*) were implicated in lifetime cannabis use. The only gene to overlap across gene-based and TWAS analyses was *CADM2* (**Supplementary Table 10**). Of the 44 unique genes associated with lifetime and frequency of cannabis use, 29 gene associations have not been previously associated with any cannabis-related trait (**Supplementary Table 10**).

Gene-set and tissue-based enrichment analyses yielded no significant results (**Supplementary Tables 11-12**).

### Lifetime and Frequency of Cannabis Use Are Genetically Correlated with Psychiatric, Cognitive, and Physical Health Traits

There were 115 traits genetically correlated (***r_g_***) with lifetime cannabis use and 38 with frequency of cannabis use after applying a 5% false discovery rate (**FDR**) correction (Fig. 2**-3**, **Supplementary Table 13**). We identified 29 traits that were significantly genetically correlated with both lifetime and frequency of cannabis use (10 anthropomorphic traits; 19 psychiatric traits), which were usually consistent in their direction of effect, with exceptions for intelligence and executive function (positively genetically correlated with frequency of use, negatively genetically correlated with lifetime use), and tense/‘highly strung’ and delay discounting (negatively genetically correlated with frequency of use, positively genetically correlated with lifetime use), as we review below (**Supplementary** Fig. 5).

**Figure 2.**
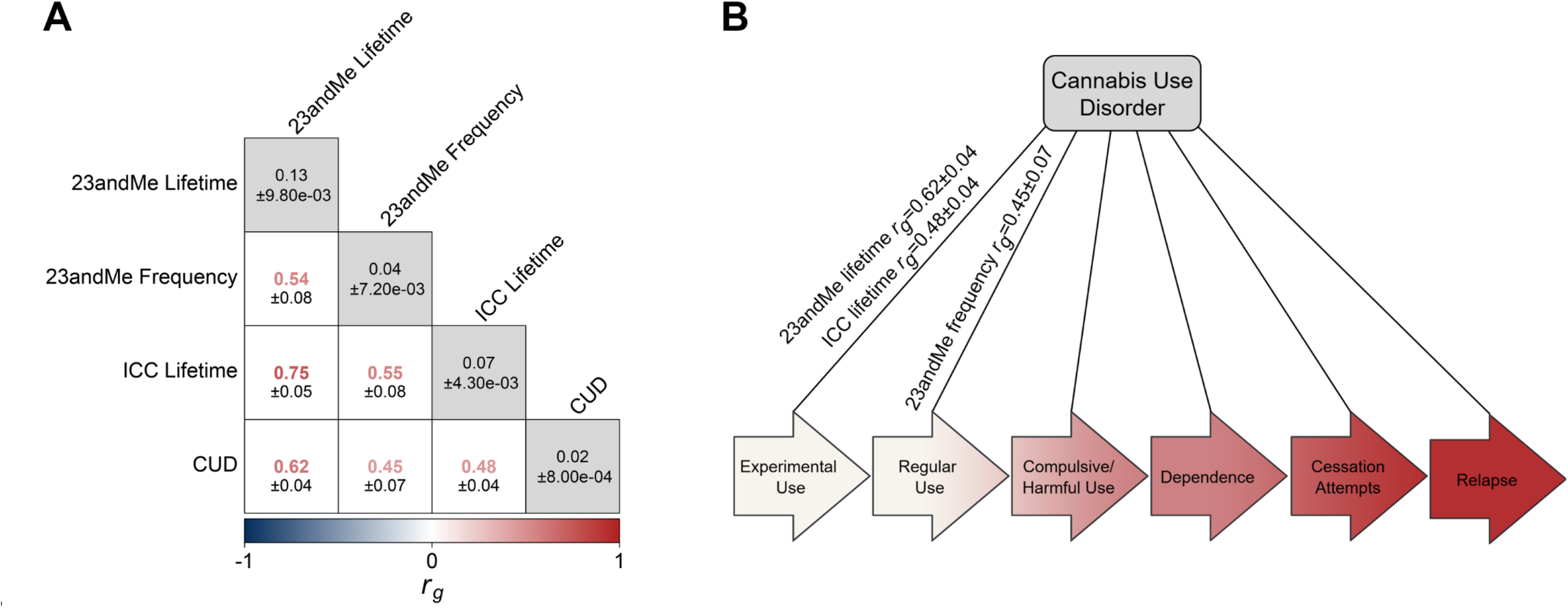
SNP-based heritability and genetic correlation analysis comparisons across cannabis-related traits. **A)** Genetic correlations and *h_2_*_SNP_ across 23andMe lifetime cannabis use and frequency of cannabis use with ICC lifetime cannabis use ^34^ and CUD from Levey *et al.* ^22^. *h_2_*_SNP_±standard error shown in matrix diagonal (gray boxes), *r_g_*±standard error in off-diagonal (white boxes). Correlation coefficients shown in heatmap color, with *p* value underneath in black. **B)** CUD requires progression through multiple pre-addiction stages, including experimental use, regular use, compulsive/harmful use, dependence, cessation attempts, and relapse. Aside from lifetime cannabis use as a proxy for experimental use and frequency of cannabis use as a proxy for regular use, which positively genetically correlate with CUD, most of these stages have not been genetically explored with GWAS.

**Figure 3.**
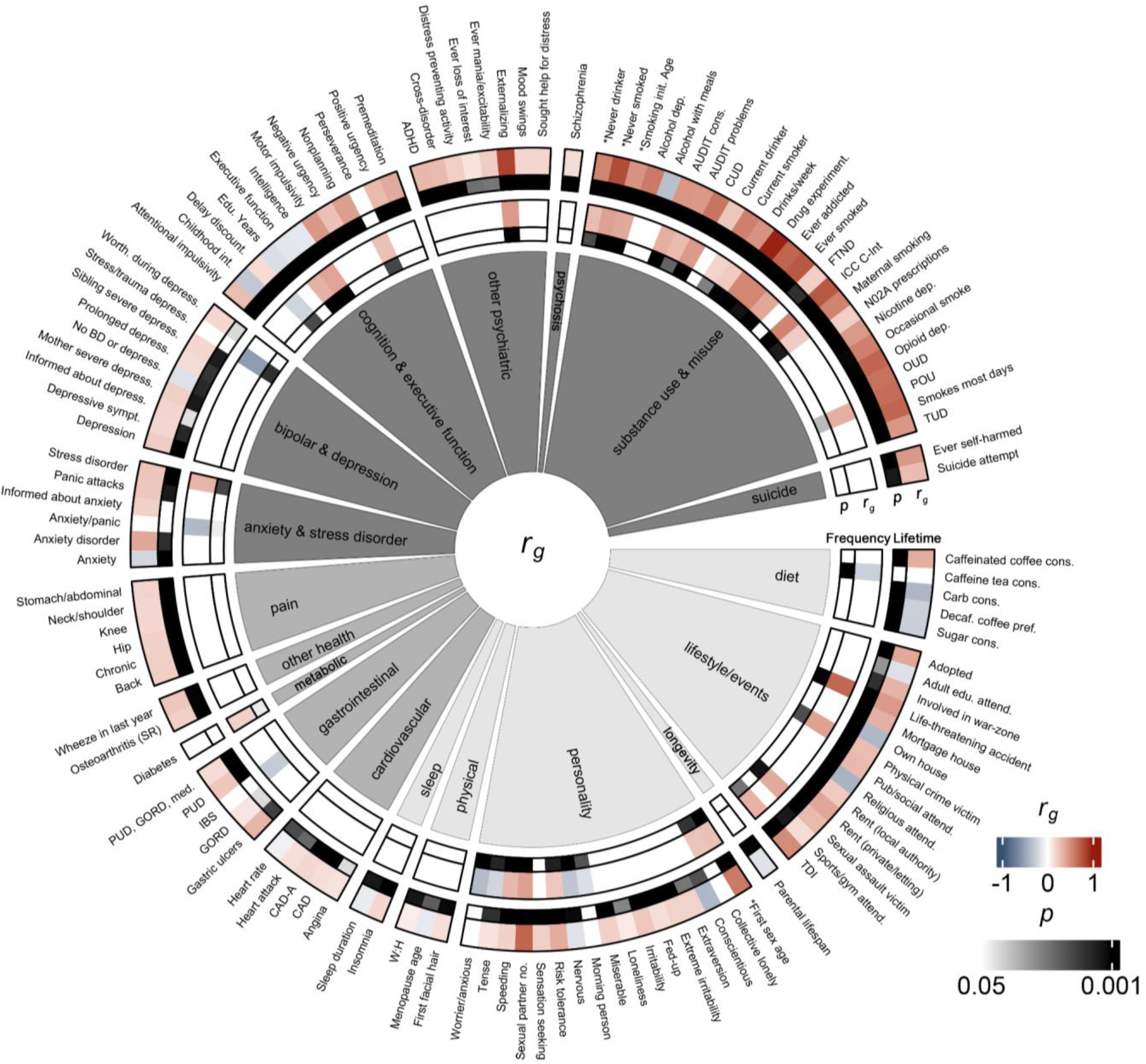
Comparison of genetic correlations across anthropometric (light gray), health (medium gray), and psychiatric (dark gray) traits between lifetime cannabis use (lanes 1 and 2) and frequency of cannabis use (lanes 3 and 4). Lanes 1 and 3 show *r_g_* values calculated by LDSC, and lanes 2 and 4 show FDR-corrected *p* values. Only traits for which at least one cohort was FDR-significant are displayed. For a full list of correlations and trait names, see **Supplementary Table 13**. *reverse coded traits.

#### Cannabis and Other Substance Use Traits

The genetic correlation between lifetime and frequency of cannabis use was moderate (*r_g_*=0.54±0.08, *p*=1.89E-10), suggesting imperfect genetic overlap between the two traits. We identified positive genetic correlations between CUD and lifetime (*r_g_*=0.62±0.04, *p*=2.44E-59), as well as frequency of cannabis use (*r_g_*=0.45±0.07, *p*=2.45E-10; Fig. 2). Compared to lifetime cannabis use from the ICC, our lifetime cannabis use trait was more strongly genetically correlated with CUD (23andMe-CUD *r_g_*=0.62±0.04, *p*=2.44E-59 vs. ICC-CUD *r_g_*=0.48±0.04, *p*=4.30E-33). Positive genetic correlations with other aspects of substance use (e.g., drug experimentation and lifetime cannabis use: *r_g_*=0.97±0.01, *p*<1.35E-161; frequency: *r_g_*=0.54±0.07, *p*=5.45E-14) and misuse (e.g., Alcohol Use Disorder Identification Test (**AUDIT**) problems and lifetime cannabis use: *r_g_*=0.46±0.06, *p*=1.54E-15; frequency of cannabis use: *r_g_*=0.30±0.10, *p*=2.46E-03) were among the top genetic correlations for lifetime and frequency of cannabis use (Fig. 3, **Supplementary Table 13**).

#### Psychiatric Disorders

Lifetime cannabis use was genetically correlated with schizophrenia (*r_g_*=0.15±0.03, *p*=7.33E-07); however, frequency of cannabis use was not (*r_g_*=0.02±0.05, *p*=0.73). We also identified associations with other psychiatric traits and lifetime cannabis use like ADHD (*r_g_*=0.31±0.05, *p*=5.20E-12), depression (*r_g_*=0.22±0.04, *p*=3.52E-10), and cross-disorder (*r_g_*=0.30±0.05, *p*=3.91E-10). We identified significant genetic correlations between frequency of cannabis use and the psychiatric-related traits “depression possibly related to stressful or traumatic events” (*r_g_*=-0.54±0.16, *p*=9.22E-04), stress-related disorder (*r_g_*=0.33±0.10, *p*=1.44E-03), and anxiety/panic attacks (*r_g_*=-0.38±0.14, *p*=6.06E-03), though only stress-related disorder was also genetically correlated with lifetime cannabis use (*r_g_*=0.25±0.06, *p*=1.42E-04).

#### Externalizing and Risk-Taking Traits

Among the strongest associations for lifetime cannabis use were positive genetic correlations with externalizing behavior (*r_g_*=0.84±0.03, *p*=5.65E-208), and two traits that were used to construct externalizing behavior^44^: number of sexual partners (*r_g_*=0.69±0.03, *p*=6.16E-115) and age at first sex (reverse-coded; *r_g_*=0.60±0.03, *p*=1.08E-83). We found similar positive genetic correlations with frequency of cannabis use and externalizing and risk-taking (externalizing: *r_g_*=0.45±0.06, *p*=1.68E-15); number of sexual partners: *r_g_*=0.42±0.06, *p*=3.17E-12).

#### Cognitive Traits

We identified significant genetic correlations between lifetime cannabis use and 11 cognitive and executive function-related traits; these included positive genetic correlations with delay discounting (*r_g_*=0.16±0.04, *p*=3.51E-04) and other impulsivity-related measures (*r_g_*=0.27±0.05 to 0.46±0.05, *p*=1.02E-22 to 3.20E-04), and negative genetic correlations with childhood intelligence (*r_g_*=-0.29±0.08, *p*=3.20E-04), educational years (*r_g_*=-0.17±0.03, *p*=1.84E-07), common executive function (*r_g_*=-0.13±0.03, *p*=3.63E-05), and intelligence (*r_g_*=-0.12±0.03, *p*=3.04E-05).

For frequency of cannabis use, we identified positive genetic correlations with intelligence (*r_g_*=0.40±0.05, *p*=4.18E-14) and common executive function (*r_g_*=0.34±0.06, *p*=7.86E-09). There was also a negative genetic correlation with delay discounting (*r_g_*=-0.23±0.07, *p*=1.62E-03), indicating those who use cannabis more frequently may devalue delayed rewards. Consistent with lifetime cannabis use, we found positive genetic correlation with the impulsivity-related measure perseverance (*r_g_*=0.28±0.09, *p*=1.48E-03).

#### Physical Health Traits

We identified genetic correlations between lifetime cannabis use and 17 physical health traits, including chronic pain (*r_g_*=0.21±0.04, *p*=5.59E-09), back pain (*r_g_*=0.22±0.05, *p*=2.19E-06), and coronary artery disease with angina (*r_g_*=0.17±0.04, *p*=2.59E-05). For frequency of cannabis use, there was a positive genetic correlation with diabetes (*r_g_*=0.20±0.07, *p*=5.96E-03) and a negative genetic correlation with irritable bowel syndrome (*r_g_*=-0.27±0.10, *p*=6.55E-03).

### Lifetime and Frequency of Cannabis Use Polygenic Scores Associate with Cannabis Use Phenotypes

Lifetime and frequency PGS associations with cannabis use traits in *All of Us* (**AoU**) were considered in single (i.e., models only incorporating lifetime *or* frequency of cannabis use PGS as variables) and joint (i.e., models incorporating lifetime *and* frequency of cannabis use PGS as variables) PGS models (**Supplementary Tables 14-16**). In the joint-PGS model simultaneously accounting for lifetime and frequency PGS in the European cohort, based on genetic similarity (see **Methods**), lifetime cannabis use PGS associated with lifetime cannabis use (β=0.19±0.01, *p*<2.00E-16), daily cannabis use (β=0.09±0.03, *p*=5.09E-04), and problematic cannabis use (β=0.22±0.02, *p*<2.00E-16; **Table 1, Supplementary Table 16**). Frequency of cannabis use PGS was associated with lifetime cannabis use (β=0.06±0.01, *p*<2.00E-16), and nominally associated with problematic cannabis use (β=0.06±0.03, *p*=0.01), which did not survive multiple testing correction. Lifetime and frequency PGSs were estimated to explain 0.31-1.52% of the phenotypic variance in cannabis use traits (Fig. 4). In the African cohort, based on genetic similarity (see **Methods**), lifetime cannabis use was predicted by the lifetime PGS (β=0.08±0.01, *p*=2.76E-12) and the frequency PGS (β=0.04±0.01, *p*=1.88E-04), which contributed an estimated 0.20% to phenotypic variance. In both populations, phenotypic variance was primarily attributable to the lifetime cannabis use PGS versus the frequency of cannabis use PGS.

**Figure 4.**
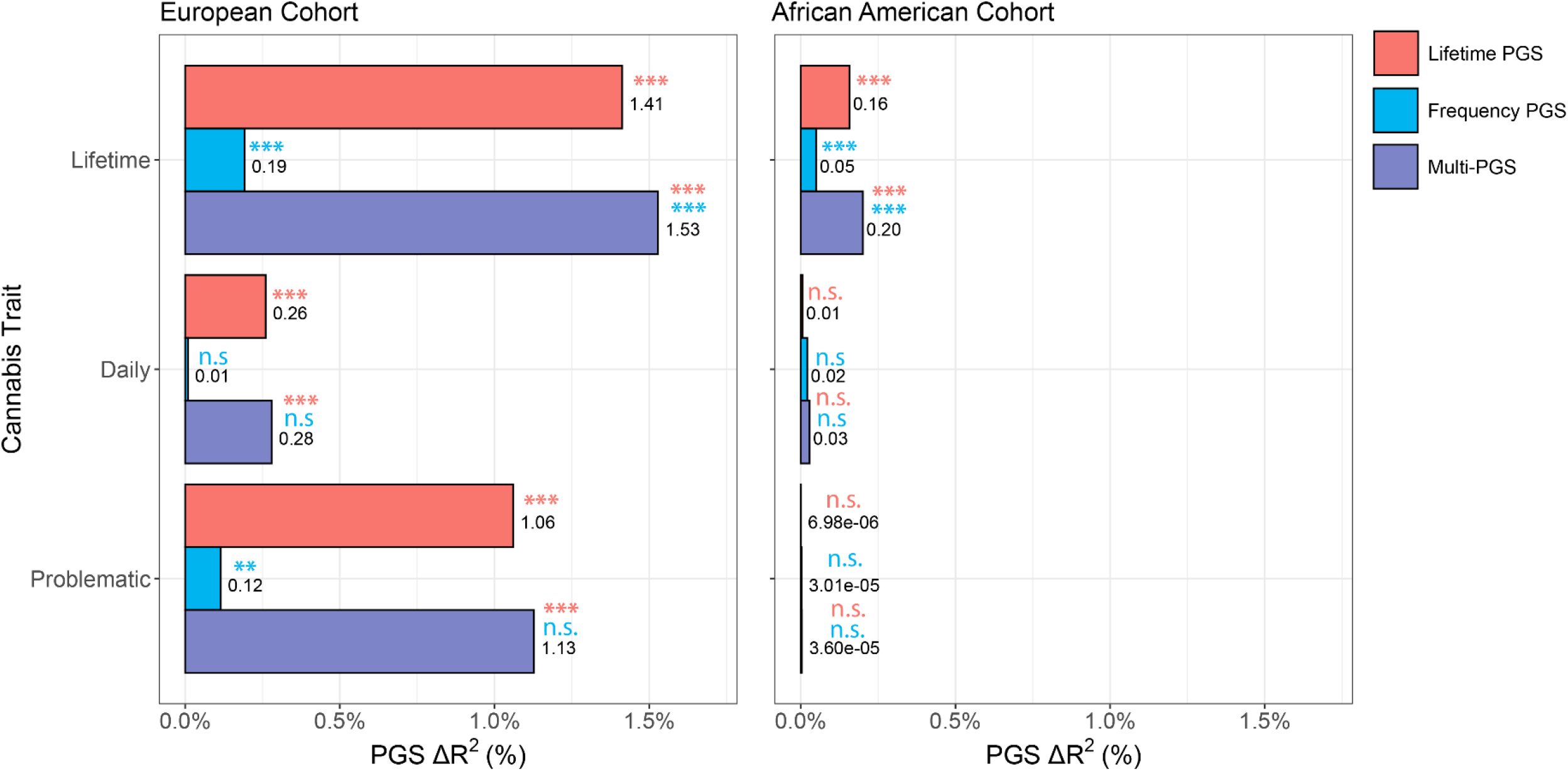
Percent proportion of lifetime, daily, and problematic cannabis use variance attributable to lifetime cannabis use PGS, frequency of cannabis use PGS, or both (joint-PGS) in European and African AoU cohorts. Bonferroni-corrected significance of PGS contribution for single- and joint-PGS models (see **Table 1, Supplementary Tables 15-16**) shown above data label in its corresponding legend color (n.s. *p*>0.05, \**p*<0.05, \*\**p*<0.01, **p<0.001).

**Table 1.** Joint-PGS regression analysis associating lifetime cannabis use PGS, frequency of cannabis use PGS, and select covariates with lifetime, daily, and problematic cannabis use in AoU cohorts. Bold PGS results are significant following Bonferroni correction (*p*<8.33E-03). For full analysis variables, see **Supplementary Table 16**.

|  | European Cohort |  |  |  |  |  |  |  |  | African Cohort |  |  |  |  |  |  |  |  |
| --- | --- | --- | --- | --- | --- | --- | --- | --- | --- | --- | --- | --- | --- | --- | --- | --- | --- | --- |
| | Lifetime Use<br>($N_{\text{case}}=64,711$,<br>$N_{\text{control}}=49,595$) | | | Daily Use<br>($N_{\text{case}}=1,411$,<br>$N_{\text{control}}=28,112$) | | | Problematic Use<br>($N_{\text{case}}=1,825$,<br>$N_{\text{control}}=118,704$) | | | Lifetime Use<br>($N_{\text{case}}=26,064$,<br>$N_{\text{control}}=21,610$) | | | Daily Use<br>($N_{\text{case}}=2,483$,<br>$N_{\text{control}}=11,718$) | | | Problematic Use<br>($N_{\text{case}}=2,315$,<br>$N_{\text{control}}=50,262$) | | |
| Variable | $\beta$ | StdErr | $p$ | $\beta$ | StdErr | $p$ | $\beta$ | StdErr | $p$ | $\beta$ | StdErr | $p$ | $\beta$ | StdErr | $p$ | $\beta$ | StdErr | $p$ |
| Lifetime PGS | **0.19** | **0.01** | **<2.00E-16** | **0.09** | **0.03** | **5.09E-04** | **0.22** | **0.02** | **<2.00E-16** | **0.08** | **0.01** | **2.76E-12** | -0.02 | 0.03 | 0.52 | 3.62E-03 | 0.02 | 0.88 |
| Frequency PGS | **0.06** | **0.01** | **<2.00E-16** | -0.03 | 0.03 | 0.38 | 0.06 | 0.03 | 0.01 | **0.04** | **0.01** | **1.88E-04** | 0.03 | 0.03 | 0.23 | 0.01 | 0.03 | 0.61 |

In all models, age was a significant negative predictor and being a male was a significant positive predictor of problematic, daily, and lifetime cannabis use (**Supplementary Tables 14-17**).

### Lifetime Cannabis Use Polygenic Score Associates with Psychiatric and Infectious Disease Diagnoses

Our phenome- and laboratory-wide association studies (**PheWAS/LabWAS**) uncovered 15 FDR-significant PheWAS associations and 9 FDR-significant LabWAS associations with lifetime cannabis use in the BioVU European cohort, as described below (Fig. 5; **Supplementary Tables 19-20**). When CUD was included as a covariate, 8 PheWAS and 6 LabWAS associations remained, as we discuss below. Tobacco smoking is prevalent among cannabis users^45^; 4 PheWAS and 4 LabWAS associations persisted when adjusting for tobacco use disorder (**TUD**), and 1 PheWAS and 5 LabWAS associations persisted when CUD and TUD were jointly included as covariates, described further below. We found no significant associations with cannabis use frequency in the European cohort, nor any significant associations for lifetime or frequency of cannabis use in the African cohort.

**Figure 5.**
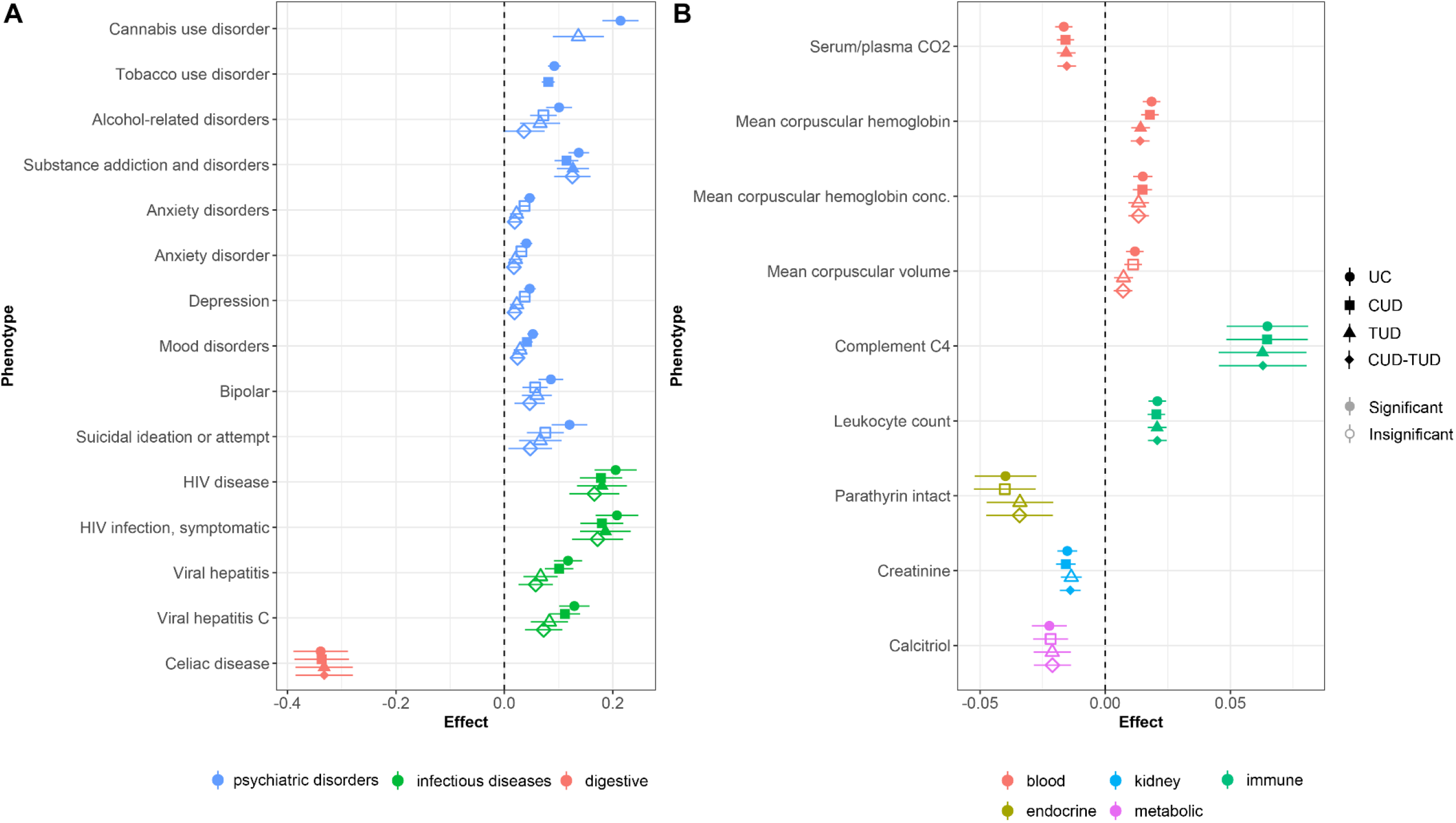
Forest plot of FDR-significant phenome associations with lifetime cannabis use PGS unconditioned (**UC**), or with adjustment for cannabis use disorder (**CUD**), tobacco use disorder (**TUD**), or both (**CUD-TUD**). **A)** PheWAS results. **B)** LabWAS results. For full trait information, see **Supplementary Tables 19-20**.

#### Psychiatric Disorders

Our PheWAS identified positive associations between lifetime cannabis use PGS and seven psychiatric disorders: TUD (β=0.09±0.01, *p*=2.44E-15), substance addiction and disorders (β=0.14±0.02, *p*=8.56E-13), CUD (β=0.21±0.03, *p*=1.24E-10), alcohol-related disorders (β=0.10±0.02, *p*=2.43E-05), mood disorder (β=0.05±0.01, *p*=3.38E-07), two anxiety traits (anxiety disorders: β=0.05±0.01, *p*=8.85E-06; anxiety disorder: β=0.04±0.01, *p*=2.55E-04), depression (β=0.05±0.01, *p*=1.73E-05), bipolar (β=0.09±0.02, *p*=1.59E-04), and suicide ideation or attempt (β=0.12±0.03, *p*=2.64E-04). TUD, substance addiction and disorders, and mood disorders persisted following adjustment for CUD, only substance addiction and disorders persisted following control for TUD, and no psychiatric disorders were significant following control for both CUD and TUD. We did not find evidence of an association with schizophrenia (β=0.02±0.06, *p*=0.68), schizophrenia and other psychotic disorders (β=0.03±0.03, *p*=0.29), or psychosis (β=0.08±0.04, *p*=0.07).

#### Infectious Diseases

We found significant positive associations between lifetime cannabis use and infectious diseases, including human immunodeficiency virus (**HIV**) disease (β=0.21±0.04, *p*=1.14E-07), symptomatic HIV infection (β=0.21±0.04, *p*=1.26E-07), viral hepatitis C (β=0.13±0.03, *p*=3.99E-06), and viral hepatitis (β=0.11±0.03, *p*=6.17E-06). All associations persisted following control for CUD, both HIV associations persisted following control for TUD, but no infectious disease associations persisted following control for both CUD and TUD.

#### Other Diagnoses

Lifetime cannabis use PGS was negatively associated with one digestive trait, celiac disease (β=-0.34±0.05, *p*=1.55E-11). This association persisted with following control for CUD, TUD, and combined CUD and TUD.

#### Blood Laboratory Biomarkers

LabWAS revealed associations with lifetime cannabis use across four blood biomarkers: mean corpuscular hemoglobin (**MCH**; β=0.02±3.53E-03, *p*=1.60E-07), carbon dioxide serum/plasma (β=-0.02±3.47E-03, *p*=1.92E-06), MCH concentration (β=0.02±3.85E-03, *p*=9.41E-05), and mean corpuscular volume (β=0.01±3.53E-03, *p*=7.77E-04). Following CUD adjustment, all but mean corpuscular volume remained significant; following adjustment for TUD alone or alongside CUD, carbon dioxide serum/plasma and MCH remained significant.

#### Immune Laboratory Biomarkers

Two immune biomarkers, leukocytes in blood (β=0.02±3.51E-03SE, *p*=2.778E-09) and complement C4 in serum or plasma (β=0.06±0.02, *p*=6.84E-05), were positively associated with lifetime cannabis use. Both remained significant following control with TUD and CUD independently or together.

#### Other Laboratory Biomarkers

The kidney biomarker creatinine in blood (β=-0.02±3.90E-03, *p*=1.02E-04), endocrine biomarker parathyrin intact in serum or plasma (β=-0.04±0.01, *p*=1.25E-03), and the metabolic biomarker calcitriol in serum and plasma (β=-0.02±0.01, *p*=1.37E-03) were negatively associated with lifetime cannabis use; none were significant following control for TUD, but creatinine in blood remained significant when CUD, and when CUD and TUD were used as covariates.

## Discussion

This study contributes to the growing body of cannabis use genetics literature by providing new GWASs of 131,895 individuals of European genetic similarity assessed for lifetime cannabis use and, for the first time, 73,374 individuals assessed for frequency of cannabis use. Both GWASs replicated the robust associations with variants nearby *CADM2* and lifetime cannabis use GWAS identified one novel locus near *GRM3*. We found that lifetime and frequency of cannabis use reliably genetically correlated with substance use-related traits, including CUD, and PGSs for both traits associated with cannabis use phenotypes in AoU. Polygenic analysis of lifetime cannabis use also revealed positive associations with substance use and mood disorders consistent with the literature, and novel phenotypic associations with anxiety disorders, infectious diseases, and red blood cell biomarkers. Overall, these results support the value of cannabis use phenotypes spanning the addiction spectrum in the exploration of genetic factors influencing cannabis use vulnerability and health risk.

Pre-addiction phenotypes are intended to capture prodromal symptoms of SUD; lifetime and frequency of cannabis use are heritable risk factors for CUD development^15,27,29,30,32,34^. They can be easily self-reported in large cohorts, making them attractive targets for GWAS. Lifetime cannabis use captures both experimental/occasional and heavy use; despite the simplicity of this phenotype, we uncovered multiple novel genetic associations with lifetime cannabis use (i.e., *GRM3* locus, genetic correlations, polygenic associations), and found it reliably associated with CUD and multiple other important traits. Although frequency of use may better account for regular cannabis use, this trait did not associate with CUD to a greater degree compared to lifetime cannabis use (*r_g_*=0.45±0.07 vs. 0.62±0.04), potentially due to lower power (*N=*73,374 vs. 131,895). However, lifetime and frequency of cannabis use was genetically correlated with each other and their associations with other complex traits were almost always directionally consistent. This included positive genetic correlations between lifetime and frequency of use with other substance use, misuse, and behavioral traits thought to be substance use risk factors like externalizing, impulsivity, and risk-taking^21,44,46–50^, consistent with ICC lifetime cannabis use genetic correlations^34^ and reports of a general “addiction risk factor” or externalizing factor accounting for genetic overlap across substances^23,44,51–53^. We previously demonstrated that consumption and problematic use phenotypes (i.e., alcohol^24,54,55^, tobacco^56^) are correlated but non-identical traits; this is likely true for cannabis. Future multivariate analyses incorporating lifetime, frequency, and other cannabis use GWASs (e.g., CUD, dependence, craving, etc.) could effectively boost locus discovery, identify novel relationships between CUD behaviors and health, and parse genomic factors pertaining to the stages of CUD^36^, as we and others have previously demonstrated for other substance use traits^23,54,55,57–59^.

One of our most notable findings was a novel association between lifetime cannabis use and rs12673181, which is located upstream of the *GRM3* gene that encodes the group II inhibitory glutamate receptor mGlu_3_. There are no known associations with this or other *GRM3* SNPs with cannabis-related traits, and while GWASs implicate *GRM3* variants in other substance use (i.e., alcohol, smoking)^60^, schizophrenia^61–65^, neuroticism^66,67^, educational attainment^68^, and other phenotypes^69–71^, these variants are not in LD with rs12673181. Recent studies also suggest that mGlu_3_ potentiates activity of mGlu ^72^, which has also garnered attention for its potential role in addictive-like behaviors and endocannabinoid synthesis^73,74^. While rs12673181 lies upstream of *GRM3,* it is not a known expression quantitative trait locus (**eQTL**) of *GRM3* (**Supplementary Table 5**)^75^. Further functional work, especially pertaining to the regulation of *GRM3*, is required to characterize its association with cannabis use vulnerability.

Through multiple lines of evidence, we found lifetime and frequency of cannabis use associated with the *CADM2* gene, replicating prior GWASs of lifetime cannabis use and CUD^23,34^. Other GWASs have found an association between SNPs in *CADM2* and other substance use traits^23,48,51,60,76–91^, risk-taking^76,85,89,92–94^, impulsivity^48^, and externalizing behaviors^44^.

Supporting the genetic correlation observed across cannabis GWAS data, PGSs for lifetime and, to a lesser degree, frequency of cannabis use, associated with phenotypes across the CUD progression spectrum (i.e., lifetime, daily, and problematic use). More variance was explained by lifetime (0.29-1.40%) rather than frequency of use PGS (0.12-0.19%), and together they explained up to 1.6% of phenotypic variance. This is on par with recent substance use PGS analyses^95–99^, including by Hodgson et al. ^100^, who estimated that ICC lifetime cannabis use PGS predicted 0.82% of variance in lifetime cannabis use and 1.2% of variance in continued cannabis use in UK Biobank participants. Although it is improbable that cannabis use PGS alone will be sufficient for clinical utility^101^, lifetime and frequency of cannabis use PGS could be useful for models predicting problematic cannabis use risk.

Largely consistent with the genetic correlations we observed, PheWAS uncovered positive associations between lifetime cannabis use PGS with substance use, depression, anxiety, bipolar, and suicidality in the BioVU cohort (*N*<66,917). To our knowledge, the positive associations with HIV and hepatitis diagnoses, negative association with celiac disease, and mixed associations with multiple blood and immune laboratory biomarkers are novel. Our findings complement a recent PheWAS conducted in the Yale-Penn sample (*N*<10,610), which is a cohort deeply phenotyped for psychiatric disorder diagnoses and related diagnostic criteria. That study found ICC lifetime cannabis use PGS positively associated with CUD, as well as traits related to other substance use (e.g., alcohol, tobacco, sedatives, stimulants) and depression^102^. That many of these relationships disappear when controlling for CUD in our PheWAS and in the Kember *et al.* ^102^ study, as well as when controlling for TUD in our study, supports the hypothesis that these associations are mediated by regular cannabis and tobacco use rather than genetic liability for lifetime cannabis use. Furthermore, like others^102^, we found minimal evidence of a relationship between lifetime cannabis use genetics, schizophrenia, and psychosis (aside from bipolar), despite the genetic relationship between cannabis use and psychosis being the subject of intense interest^103–106^following observations of their apparent bidirectional phenotypic relationship^107^. Epidemiological evidence supports a link between cannabis and heavy or high potency cannabis use^108–110^. Identifying more robust variant associations, especially for frequency of cannabis use, will aid future causal inference analyses that can resolve the role of cannabis genetics in health.

Our results were consistent with the prior cannabis use GWAS by ICC^34^. Lifetime cannabis use measured in US-based 23andMe research participants was genetically correlated with the same trait examined in the ICC cohort, which is composed of participants across North America, Europe, and Australia^34^. Both lifetime cannabis use datasets were genetically correlated with CUD, but the magnitude of this association was stronger in the 23andMe dataset compared to ICC (*r_g_*=0.62 vs. 0.48) despite our smaller sample size. Heritability estimates for our lifetime cannabis use trait was also higher (12.88% vs. 6.63%). Heritability may decrease when meta-analyzing cohorts, possibly due to cohort-specific environmental/geocultural differences that could exist surrounding cannabis use^111–113^. Furthermore, while we found consistent positive correlations with psychiatric disorders, including schizophrenia^21,34,49^, ADHD^21,34,47,49^, bipolar disorder^34,49^, and depression^21,49^ between 23andMe and ICC lifetime cannabis use, we also observed that the genetic correlation with educational attainment was negative with 23andMe and positive with ICC lifetime cannabis use^34^. Interestingly, while most genetic correlations between lifetime and frequency of cannabis use were also mostly in agreement, lifetime cannabis use negatively genetically correlated with intelligence and common executive function and positively genetically correlated with delay discounting, while we saw the inverse with frequency of use. This is not entirely unprecedented, as relationships between cannabis use and cognitive traits can be paradoxical, especially among those with psychiatric disorders, such as those with psychosis who use cannabis exhibiting greater cognitive abilities than those who do not^114^. In sum, although most associations were consistent, the differences we observed in trait heritability and patterns of genetic correlations suggest some disunity between 23andMe and ICC lifetime cannabis use cohorts, as well as lifetime and frequency of cannabis use data, which will warrant careful consideration before attempting to meta-analyzing GWAS data.

There are several limitations to our study. The legal status of cannabis use differs across countries and even US states and has been changing over the last several decades. Thus, for some of our older subjects, both lifetime and frequency of use could be reflecting use decades ago, whereas others are referencing more recent use. Most studies suggest that legalizing recreational cannabis use increases lifetime and frequency of use rates^115^, which may have impacted our findings in complex ways that depend on which location a given participant was in at the time of their use. In addition, frequency of cannabis determined by the number of use days over a 30-day window does not accurately reflect lifetime use intensity because it does not account for the duration of regular use or use quantity. These characteristics are important to CUD trajectory and other health and wellbeing relationships^116–119^. Lifetime and frequency of cannabis use GWASs also relied on self-reported data. Cannabis use is most common during adolescence and young adulthood^120^, but participants in this study averaged in their 50s and could have been at greater risk for recall bias regarding cannabis use in early life^121^. Socioeconomic variables are also associated with cannabis use rates^122,123^, and the on-average higher socioeconomic status of 23andMe research participants may have influenced our findings^36^. Finally, GWASs were conducted using genomic information from individuals of genetically predicted European ancestry. While we extended our polygenic analyses to African cohorts, cross-population transferability of PGS is suboptimal compared to investigations where discovery and target populations are ancestrally aligned^124,125^. This, along with lower sample numbers, may explain why we observed fewer associations in African versus European cohorts. Due to sample size constraints, we also did not explore associations in other ancestral groups, further limiting the generalizability of our results.

This project showcases the utility of pre-addiction phenotypes in cannabis use genomic discovery. Lifetime and frequency of cannabis use genetically associated with CUD and other SUD, alongside concerning health and psychiatric problems. Increasing sample size and investigating other heritable, diverse phenotypes (e.g., drug responsivity, craving, withdrawal; Figure 2B) will be integral to further our understanding of CUD vulnerability and the health consequences of cannabis use.

## Data Availability

We will provide 23andMe summary statistics for the top 10,000 SNPs upon publication. 23andMe GWAS summary statistics will be made available through 23andMe to qualified researchers under an agreement with 23andMe that protects the privacy of the 23andMe participants. Please visit https://research.23andme.com/collaborate/#dataset-access/ for more information and to apply to access the data. We will share the Jupyter notebooks used for PGS analysis in AoU with registered All of Us researchers upon request.

## Acknowledgements

We would like to thank the research participants and employees of 23andMe for making this work possible. Participants provided informed consent and volunteered to participate in the research online, under a protocol approved by the external AAHRPP-accredited Institutional Review Board (**IRB**), Ethical & Independent (**E&I**) Review Services. As of 2022, E&I Review Services is part of Salus IRB (https://www.versiticlinicaltrials.org/salusirb). The following members of the 23andMe Research Team contributed to this study: Stella Aslibekyan, Adam Auton, Elizabeth Babalola, Robert K. Bell, Jessica Bielenberg, Katarzyna Bryc, Emily Bullis, Daniella Coker, Gabriel Cuellar Partida, Devika Dhamija, Sayantan Das, Teresa Filshtein, Kipper Fletez-Brant, Will Freyman, Karl Heilbron, Pooja M. Gandhi, Karl Heilbron, Barry Hicks, David A. Hinds, Ethan M. Jewett, Yunxuan Jiang, Katelyn Kukar, Keng-Han Lin, Maya Lowe, Jey C. McCreight, Matthew H. McIntyre, Steven J. Micheletti, Meghan E. Moreno, Joanna L. Mountain, Priyanka Nandakumar, Elizabeth S. Noblin, Jared O’Connell, Aaron A. Petrakovitz, G. David Poznik, Morgan Schumacher, Anjali J. Shastri, Janie F. Shelton, Jingchunzi Shi, Suyash Shringarpure, Vinh Tran, Joyce Y. Tung, Xin Wang, Wei Wang, Catherine H. Weldon, Peter Wilton, Alejandro Hernandez, Corinna Wong, Christophe Toukam Tchakouté.

We would also like to thank The Externalizing Consortium for sharing the GWAS summary statistics of externalizing. The Externalizing Consortium: Principal Investigators: Danielle M. Dick, Philipp Koellinger, K. Paige Harden, Abraham A. Palmer. Lead Analysts: Richard Karlsson Linnér, Travis T. Mallard, Peter B. Barr, Sandra Sanchez-Roige. Significant Contributors: Irwin D. Waldman. The Externalizing Consortium has been supported by the National Institute on Alcohol Abuse and Alcoholism (R01AA015416 -administrative supplement), and the National Institute on Drug Abuse (R01DA050721). Additional funding for investigator effort has been provided by K02AA018755, U10AA008401, P50AA022537, as well as a European Research Council Consolidator Grant (647648 EdGe to Koellinger). The content is solely the responsibility of the authors and does not necessarily represent the official views of the above funding bodies. The Externalizing Consortium would like to thank the following groups for making the research possible: 23andMe, Add Health, Vanderbilt University Medical Center’s BioVU, Collaborative Study on the Genetics of Alcoholism (COGA), the Psychiatric Genomics Consortium’s Substance Use Disorders working group, UK10K Consortium, UK Biobank, and Philadelphia Neurodevelopmental Cohort.

We gratefully acknowledge *All of Us* participants for their contributions, without whom this research would not have been possible. We also thank the National Institutes of Health’s *All of Us* Research Program for making available the participant data examined in this study. The *All of Us* Research Program is supported by the National Institutes of Health, Office of the Director: Regional Medical Centers: 1 OT2 OD026549; 1 OT2 OD026554; 1 OT2 OD026557; 1 OT2 OD026556; 1 OT2 OD026550; 1 OT2 OD 026552; 1 OT2 OD026553; 1 OT2 OD026548; 1 OT2 OD026551; 1 OT2 OD026555; IAA #: AOD 16037; Federally Qualified Health Centers: HHSN 263201600085U; Data and Research Center: 5 U2C OD023196; Biobank: 1 U24 OD023121; The Participant Center: U24 OD023176; Participant Technology Systems Center: 1 U24 OD023163; Communications and Engagement: 3 OT2 OD023205; 3 OT2 OD023206; and Community Partners: 1 OT2 OD025277; 3 OT2 OD025315; 1 OT2 OD025337; 1 OT2 OD025276.

## Author Contributions

SSR and AAP conceived the idea. PF and SLE contributed formal analyses and curation of 23andMe data. HHAT contributed to formal analyses, investigation, and data visualization. contributed to formal data analysis and data visualization. JJM, MVJ, RBC, and SP contributed to formal analyses. HHAT and SSR wrote the manuscript. All authors reviewed and edited the manuscript.

## Methods

### Participants and GWASs

Lifetime and frequency of cannabis use GWASs were conducted in male and female 23andMe research participants of European genetic similarity, as previously described^48^. Ancestry falls along a spectrum^126,127^; individuals were only included in the analysis if they had >97% European genetic similarity (see **Supplementary Methods**), as determined through local ancestry analysis^128^. Participants provided informed consent and volunteered to participate in research online under a protocol approved by the external AAHRPP-accredited Institutional Review Board (**IRB**), Ethical & Independent (**E&I**) Review Services. As of 2022, E&I Review Services is part of Salus IRB (https://www.versiticlinicaltrials.org/salusirb). During 4 months in 2015 and 14 months between 2018 to 2020, participants completed a questionnaire surveying a range of personal and behavior characteristics. Included in this survey were questions on lifetime substance use and substance use frequency. Specifically, “Yes” or “No” responses to the question “Have you ever in your life used marijuana?” were collected as a measure of lifetime cannabis use. If participants answered “Yes”, they were prompted to answer the question “How many days did you use marijuana during your heaviest 30 days?” as a measure of frequency of cannabis use. Participants could respond between 0 and 30 days.

For lifetime cannabis use and frequency of cannabis use, 23andMe conducted GWASs of up to 33,419,581 imputed genetic variants using linear regression and assuming an additive genetic model. Samples were genotyped on one of five genotyping platforms. The V1 and V2 platforms were variants of the Illumina HumanHap550 + BeadChip, including about 25,000 custom SNPs selected by 23andMe, with a total of ∼560,000 SNPs. The V3 platform was based on the Illumina OmniExpress + BeadChip, with custom content to improve the overlap with our V2 array, with a total of ∼950,000 SNPs. The V4 platform is a fully custom array, including a lower redundancy subset of V2 and V3 SNPs with additional coverage of lower-frequency coding variation, and ∼570,000 SNPs. The v5 platform, in current use, is an Illumina Infinium Global Screening Array (∼640,000 variants) supplemented with ∼50,000 variants of custom content. Samples that failed to reach 98.5% call rate were excluded from the study. We excluded SNPs of low genotyping quality, including those that failed a Mendelian transmission test in trios or with large allele frequency discrepancies compared to European 1000 Genomes reference data, failed Hardy-Weinberg Equilibrium testing, failed batch effects testing, or had a call rate <90%, as well as SNPs with a minor allele frequency<0.1% and imputed variants with low imputation quality or with evidence of batch effects (**Supplementary Table 3**). Model covariates included age, sex, the first 5 genetic principal components (**PCs**), and indicator variables for genotype platforms (see **Supplementary Methods** for additional details). Unrelated participants categorized as of European ancestry were included in the GWASs (lifetime cannabis use *N*=131,895; frequency of cannabis use *N*=73,374; Durand et al., 2014). For full details on genotyping and GWASs, see **Supplementary Methods**.

### Functional annotation and gene-Based Analyses

#### Functional annotation

Using the web-based platform Functional Mapping and Annotation of Genome-Wide Association Studies (**FUMA** v1.3.8), SNPs were annotated based on ANNOVAR categories, Combined Annotation Dependent Depletion scores, RegulomeDB scores, eQTLs, and chromatin state predicted by ChromHMM. Novel SNPs were identified as those not in LD (*r*^2^<0.10) or within ±1Mb of GWAS-significant SNPs uncovered by other GWASs of cannabis use traits (e.g., initiation, age of onset, CUD) sourced from the literature^20–23,34,35,51,129–133^ and from the EBI GWAS Catalog (https://www.ebi.ac.uk/gwas/). Novel genes were identified as those not identified by gene-based analyses in other cannabis-related studies^22,34,35,103,134–138^ or with start/stop positions within ±1Mb of previously uncovered GWAS-significant SNPs.

#### MAGMA gene-based and pathway analyses

We used Multi-marker Analysis of GenoMic Annotation (**MAGMA**, v1.08, Ensembl build v92), which is included in FUMA, to annotate SNPs to protein-coding genes. LD was estimated using the 1000 Genomes European reference sample, and significance determined following Bonferroni correction (*p<*2.53E-06). Gene-set analysis was conducted on 10,678 gene-sets and Gene Ontology terms curated from the Molecular Signatures Database (MsigDB v7.0). Tissue-specific gene expression profiles were assessed in 54 tissue types and 30 general tissue types with average gene expression in each tissue used as a covariate. Using Genome-Tissue Expression (**GTEx**, v8) RNA-seq data, gene expression values were log_2_ transformed from the average Reads Per Kilobase Million (max value=50) per tissue. Significance was determined following Bonferroni correction (*p<*9.26E-04 for 54 tissue types; *p<*1.67E-03 for 30 general tissue types).

#### H-MAGMA

We incorporated lifetime and frequency of cannabis use GWAS data with chromatin interaction profiles from human brain tissue using Hi-C coupled MAGMA (**H-MAGMA**; Sey et al., 2020). H-MAGMA assigns non-coding SNPs to genes based on chromatin interactions from fetal brain, adult brain, midbrain neurons, cortical neurons, iPSC-derived neurons, and iPSC-derived astrocytes datasets (https://github.com/thewonlab/H-MAGMA). Exonic and promoter SNPs were assigned to genes based on physical position^139^. We applied a Bonferroni correction based on the total number of gene-tissue pairs tested (*p*<9.42E-07 to 9.45E-07).

#### S-PrediXcan

We performed a transcriptome-wide association study using S-PrediXcan (v0.7.5) to identify eQTL-linked genes associated with lifetime and frequency of cannabis use^140^. S-PrediXcan uses genetic information to predict gene expression levels in various tissues and tests if eQTLs correlate with lifetime or frequency of cannabis use across 49 bodily tissues (*N_genes_*=1,619 to 9,949). S-PrediXcan uses precomputed tissue weights from the GTEx project database (https://www.gtexportal.org/) as the reference transcriptome dataset via Elastic net models. As input data, we included summary statistics, transcriptome tissue data, and covariance matrices of the SNPs within each gene model (HapMap SNP set available at the PredictDB Data Repository)^140^ from all available tissues. We applied Bonferroni correction for each tissue type (*p*<3.09E-05 to 5.03E-06).

### LDSC heritability and genetic correlations across health, psychiatric, and anthropomorphic traits

Linkage Disequilibrium Score regression (**LDSC**; https://github.com/bulik/ldsc) was used to calculate *h2_SNP_* and genetic correlations^141^. *h2_SNP_* was calculated from pre-computed LD scores (“eur_w_ld_chr/”). *r_g_* were calculated between lifetime or frequency of cannabis use with 292 other traits across 22 health, psychiatric, and lifestyle categories (**Supplementary Methods**). We applied a 5% FDR correction to account for multiple testing.

### Polygenic score analyses

#### PGS of lifetime, daily, and problematic cannabis use in AoU

We tested the associations between lifetime or frequency of cannabis use PGSs with cannabis traits available for AoU participants clustering within a European or African genetic ancestry panel (for details, see All of Us Research Program Genomics Investigators ^142^). AoU is a diverse health database currently including survey responses, physical measurements, genotyping data, and electronic health records (**EHR**) for over 400,000 individuals living in the United States^142,143^. Using survey and EHR data, participants were assigned binary identifiers for lifetime cannabis use (concept id: 1585636), daily cannabis use among those who reported cannabis use in their lifetime (concept id: 1585650), and problematic cannabis use (concept ids: 434327, 440387, 440996, 433452, 437838, 4323639, 4103419, 435231, 434019, 434328; **Supplementary Methods**).

We calculated PGSs in male or female participants who had available short-read whole genome sequencing data and applicable cannabis use data. We used the Allele Count/Allele Frequency (**ACAF**) threshold SNP callset curated by AoU, which includes SNPs of MAF > 1% or allele counts over 100 for each ancestral subpopulation. Using PRS-CS “auto” v1.1.0, the SNP set was filtered to biallelic SNPs present in the HapMap3 European ancestry set and SNPs were weighted. Lifetime and frequency of cannabis use PGSs were created from 782,975 weighted SNPs using the allelic-scoring function, *score,* in PLINK (v1.9; Ge et al., 2019). The base R function *glm* was used to fit logistic regression models for each cannabis use trait using PGS(s), as well as the additional covariates of age, sex, and the first 10 global PCs provided by AoU. Models included single PGS models (lifetime or frequency PGS + additional covariates), a joint-PGS model (lifetime PGS + frequency PGS + additional covariates), and a null model (additional covariates only). For the joint-PGS model, Bonferroni correction was applied for two tests (lifetime PGS and frequency PGS) and three outcomes (lifetime, daily, and problematic cannabis use) for a total of *N*=6 comparisons (*p*<8.33E-03); single PGS models were corrected for one test and three outcomes (N=3, *p*<1.67E-02). Joint-PGS liability scale *R*^2^ values were calculated as previously described by Lee et al. ^145^ using the *NagelkerkeR2* function in the R package *fmsb* (v0.7.6) and the estimated prevalence of cannabis use traits in US adults (**Supplementary Methods**). PGS Δ*R*^2^ was calculated by subtracting *R*^2^ calculated from models including PGS from the *R*^2^ of the null model.

#### Phenome- and Laboratory-wide association analyses in a hospital cohort (BioVU)

We tested associations between lifetime or frequency of cannabis use PGSs and medical condition liability from hospital-based cohorts using data from the Vanderbilt University Medical Center (**VUMC**; IRB #160302, #172020, #190418)^146^. The BioVU cohort, a subset of VUMC biobank participants (*N*=72,821), provided genotyping data and EHR containing clinical data and laboratory-assessed biomarkers^144,146,147^. For each unrelated European (*N*=66,917) and African (*N*=12,383) BioVU participant based on genetic similarity, we computed lifetime and frequency of cannabis use PGSs using the PRS-CS v1.1.0^144^.

For PheWAS, we fitted a logistic regression model to each case/control disease phenotypes (“phecodes”) to estimate the log odds of each diagnosis given lifetime cannabis use/frequency of cannabis use PGS, while adjusting for sex, median age of the longitudinal EHR, and the first 10 PCs with the PheWAS v0.12 R package^144^. At least two International Disease Classification (**ICD**) codes mapping to a PheWAS disease category (Phecode Map 1.2; https://phewascatalog.org/phecodes) and a minimum of 100 cases were required for phecode inclusion. We also conducted additional sensitivity analyses using TUD (phecode 318) and CUD (see **Supplementary Table 12** for CUD ICD codes) as covariates to examine if SUD mediated associations with cannabis PGSs. We calculated the 5% FDR for all associations performed (*N*=1,405).

For LabWAS, we implemented the pipeline established by Dennis *et al.* ^147^. LabWAS associates PGS with laboratory biomarkers (i.e., measurements) evaluated in BioVU participants. LabWAS uses the median, inverse normal quantile transformed age-adjusted values from the QualityLab pipeline in a linear regression to determine the association between lifetime or frequency of cannabis use PGSs with 314 phenotypes. We controlled for the same covariates as for the PheWAS analyses, excluding median age because the pipeline corrects for age using cubic splines with 4 knots. We applied 5% FDR correction across all LabWAS associations performed (*N*=314).

All results are presented as the mean±standard error unless otherwise specified.

## References

1. United Nations Office on Drug and Crime (2022). Booklet 3 - Drug market trends of Cannabis and Opioids.

2. Pacek, L.R., Mauro, P.M., and Martins, S.S. (2015). Perceived risk of regular cannabis use in the United States from 2002 to 2012: differences by sex, age, and race/ethnicity. Drug Alcohol Depend 149, 232–244. 10.1016/j.drugalcdep.2015.02.009.

3. Statistics Canada (2013). Mental and substance use disorders in Canada.

4. Rotermann, M. (2021). Looking back from 2020, how cannabis use and related behaviours changed in Canada. Health Rep 32, 3–14. 10.25318/82-003-x202100400001-eng.

5. Whiting, P.F., Wolff, R.F., Deshpande, S., Di Nisio, M., Duffy, S., Hernandez, A.V., Keurentjes, J.C., Lang, S., Misso, K., Ryder, S., et al. (2015). Cannabinoids for Medical Use: A Systematic Review and Meta-analysis. JAMA 313, 2456–2473. 10.1001/jama.2015.6358.

6. Bellocchio, L., Inchingolo, A.D., Inchingolo, A.M., Lorusso, F., Malcangi, G., Santacroce, L., Scarano, A., Bordea, I.R., Hazballa, D., D’Oria, M.T., et al. (2021). Cannabinoids Drugs and Oral Health-From Recreational Side-Effects to Medicinal Purposes: A Systematic Review. Int J Mol Sci 22. 10.3390/ijms22158329.

7. Diep, C., Tian, C., Vachhani, K., Won, C., Wijeysundera, D.N., Clarke, H., Singh, M., and Ladha, K.S. (2022). Recent cannabis use and nightly sleep duration in adults: a population analysis of the NHANES from 2005 to 2018. Reg Anesth Pain Med 47, 100–104. 10.1136/rapm-2021-103161.

8. Hasin, D.S., Kerridge, B.T., Saha, T.D., Huang, B., Pickering, R., Smith, S.M., Jung, J., Zhang, H., and Grant, B.F. (2016). Prevalence and Correlates of DSM-5 Cannabis Use Disorder, 2012-2013: Findings from the National Epidemiologic Survey on Alcohol and Related Conditions-III. Am J Psychiatry 173, 588–599. 10.1176/appi.ajp.2015.15070907.

9. Hayley, A.C., Stough, C., and Downey, L.A. (2017). DSM-5 cannabis use disorder, substance use and DSM-5 specific substance-use disorders: Evaluating comorbidity in a population-based sample. Eur Neuropsychopharmacol 27, 732–743. 10.1016/j.euroneuro.2017.06.004.

10. Howard, J., and Osborne, J. (2020). Cannabis and work: Need for more research. Am J Ind Med 63, 963–972. 10.1002/ajim.23170.

11. Keen, L., 2nd, Turner, A.D., George, L., and Lawrence, K. (2022). Cannabis use disorder severity and sleep quality among undergraduates attending a Historically Black University. Addict Behav 134, 107414. 10.1016/j.addbeh.2022.107414.

12. Lo, J.O., Hedges, J.C., and Girardi, G. (2022). Impact of cannabinoids on pregnancy, reproductive health, and offspring outcomes. Am J Obstet Gynecol 227, 571–581. 10.1016/j.ajog.2022.05.056.

13. Pacek, L.R., Herrmann, E.S., Smith, M.T., and Vandrey, R. (2017). Sleep continuity, architecture and quality among treatment-seeking cannabis users: An in-home, unattended polysomnographic study. Exp Clin Psychopharmacol 25, 295–302. 10.1037/pha0000126.

14. Page, R.L., 2nd, Allen, L.A., Kloner, R.A., Carriker, C.R., Martel, C., Morris, A.A., Piano, M.R., Rana, J.S., Saucedo, J.F., American Heart Association Clinical Pharmacology, C., et al. (2020). Medical Marijuana, Recreational Cannabis, and Cardiovascular Health: A Scientific Statement From the American Heart Association. Circulation 142, e131–e152. 10.1161/CIR.0000000000000883.

15. Feingold, D., Livne, O., Rehm, J., and Lev-Ran, S. (2020). Probability and correlates of transition from cannabis use to DSM-5 cannabis use disorder: Results from a large-scale nationally representative study. Drug Alcohol Rev 39, 142–151. 10.1111/dar.13031.

16. American Psychiatric Association (2013). Substance-Related and Addictive Disorders. In Diagnostic and Statistical Manual of Mental Disorders, (https://doi-org.subzero.lib.uoguelph.ca/10.1176/appi.books.9780890425596.dsm16).

17. Hines, L.A., Morley, K.I., Rijsdijk, F., Strang, J., Agrawal, A., Nelson, E.C., Statham, D., Martin, N.G., and Lynskey, M.T. (2018). Overlap of heritable influences between cannabis use disorder, frequency of use and opportunity to use cannabis: trivariate twin modelling and implications for genetic design. Psychol Med 48, 2786–2793. 10.1017/S0033291718000478.

18. Kendler, K.S., Ohlsson, H., Maes, H.H., Sundquist, K., Lichtenstein, P., and Sundquist, J. (2015). A population-based Swedish Twin and Sibling Study of cannabis, stimulant and sedative abuse in men. Drug Alcohol Depend 149, 49–54. 10.1016/j.drugalcdep.2015.01.016.

19. Verweij, K.J., Zietsch, B.P., Lynskey, M.T., Medland, S.E., Neale, M.C., Martin, N.G., Boomsma, D.I., and Vink, J.M. (2010). Genetic and environmental influences on cannabis use initiation and problematic use: a meta-analysis of twin studies. Addiction 105, 417–430. 10.1111/j.1360-0443.2009.02831.x.

20. Demontis, D., Rajagopal, V.M., Thorgeirsson, T.E., Als, T.D., Grove, J., Leppala, K., Gudbjartsson, D.F., Pallesen, J., Hjorthoj, C., Reginsson, G.W., et al. (2019). Genome-wide association study implicates CHRNA2 in cannabis use disorder. Nat Neurosci 22, 1066–1074. 10.1038/s41593-019-0416-1.

21. Johnson, E.C., Demontis, D., Thorgeirsson, T.E., Walters, R.K., Polimanti, R., Hatoum, A.S., Sanchez-Roige, S., Paul, S.E., Wendt, F.R., Clarke, T.K., et al. (2020). A large-scale genome-wide association study meta-analysis of cannabis use disorder. Lancet Psychiatry 7, 1032–1045. 10.1016/S2215-0366(20)30339-4.

22. Levey, D.F., Galimberti, M., Deak, J.D., Wendt, F.R., Bhattacharya, A., Koller, D., Harrington, K.M., Quaden, R., Johnson, E.C., Gupta, P., et al. (2023). Multi-ancestry genome-wide association study of cannabis use disorder yields insight into disease biology and public health implications. Nat Genet 55, 2094–2103. 10.1038/s41588-023-01563-z.

23. Xu, H., Toikumo, S., Crist, R.C., Glogowska, K., Jinwala, Z., Deak, J.D., Justice, A.C., Gelernter, J., Johnson, E.C., Kranzler, H.R., and Kember, R.L. (2023). Identifying genetic loci and phenomic associations of substance use traits: A multi-trait analysis of GWAS (MTAG) study. Addiction 118, 1942–1952. 10.1111/add.16229.

24. Sanchez-Roige, S., Palmer, A.A., and Clarke, T.K. (2020). Recent Efforts to Dissect the Genetic Basis of Alcohol Use and Abuse. Biol Psychiatry 87, 609–618. 10.1016/j.biopsych.2019.09.011.

25. McLellan, A.T., Koob, G.F., and Volkow, N.D. (2022). Preaddiction-A Missing Concept for Treating Substance Use Disorders. JAMA Psychiatry 79, 749–751. 10.1001/jamapsychiatry.2022.1652.

26. Agrawal, A., Madden, P.A., Bucholz, K.K., Heath, A.C., and Lynskey, M.T. (2014). Initial reactions to tobacco and cannabis smoking: a twin study. Addiction 109, 663–671. 10.1111/add.12449.

27. Degenhardt, L., Coffey, C., Carlin, J.B., Swift, W., Moore, E., and Patton, G.C. (2010). Outcomes of occasional cannabis use in adolescence: 10-year follow-up study in Victoria, Australia. Br J Psychiatry 196, 290–295. 10.1192/bjp.bp.108.056952.

28. Scherrer, J.F., Grant, J.D., Duncan, A.E., Sartor, C.E., Haber, J.R., Jacob, T., and Bucholz, K.K. (2009). Subjective effects to cannabis are associated with use, abuse and dependence after adjusting for genetic and environmental influences. Drug Alcohol Depend 105, 76–82. 10.1016/j.drugalcdep.2009.06.014.

29. Swift, W., Coffey, C., Carlin, J.B., Degenhardt, L., and Patton, G.C. (2008). Adolescent cannabis users at 24 years: trajectories to regular weekly use and dependence in young adulthood. Addiction 103, 1361–1370. 10.1111/j.1360-0443.2008.02246.x.

30. Windle, M., and Wiesner, M. (2004). Trajectories of marijuana use from adolescence to young adulthood: predictors and outcomes. Dev Psychopathol 16, 1007–1027. 10.1017/s0954579404040118.

31. Lyons, M.J., Toomey, R., Meyer, J.M., Green, A.I., Eisen, S.A., Goldberg, J., True, W.R., and Tsuang, M.T. (1997). How do genes influence marijuana use? The role of subjective effects. Addiction 92, 409–417.

32. Leung, J., Chan, G.C.K., Hides, L., and Hall, W.D. (2020). What is the prevalence and risk of cannabis use disorders among people who use cannabis? a systematic review and meta-analysis. Addict Behav 109, 106479. 10.1016/j.addbeh.2020.106479.

33. Haberstick, B.C., Zeiger, J.S., Corley, R.P., Hopfer, C.J., Stallings, M.C., Rhee, S.H., and Hewitt, J.K. (2011). Common and drug-specific genetic influences on subjective effects to alcohol, tobacco and marijuana use. Addiction 106, 215–224. 10.1111/j.1360-0443.2010.03129.x.

34. Pasman, J.A., Verweij, K.J.H., Gerring, Z., Stringer, S., Sanchez-Roige, S., Treur, J.L., Abdellaoui, A., Nivard, M.G., Baselmans, B.M.L., Ong, J.S., et al. (2018). GWAS of lifetime cannabis use reveals new risk loci, genetic overlap with psychiatric traits, and a causal influence of schizophrenia. Nat Neurosci 21, 1161–1170. 10.1038/s41593-018-0206-1.

35. Stringer, S., Minica, C.C., Verweij, K.J., Mbarek, H., Bernard, M., Derringer, J., van Eijk, K.R., Isen, J.D., Loukola, A., Maciejewski, D.F., et al. (2016). Genome-wide association study of lifetime cannabis use based on a large meta-analytic sample of 32 330 subjects from the International Cannabis Consortium. Transl Psychiatry 6, e769. 10.1038/tp.2016.36.

36. Sanchez-Roige, S., and Palmer, A.A. (2020). Emerging phenotyping strategies will advance our understanding of psychiatric genetics. Nat Neurosci 23, 475–480. 10.1038/s41593-020-0609-7.

37. Thorpe, H.H.A., Talhat, M.A., and Khokhar, J.Y. (2021). High genes: Genetic underpinnings of cannabis use phenotypes. Prog Neuropsychopharmacol Biol Psychiatry 106, 110164. 10.1016/j.pnpbp.2020.110164.

38. McBain, R.K., Wong, E.C., Breslau, J., Shearer, A.L., Cefalu, M.S., Roth, E., Burnam, M.A., and Collins, R.L. (2020). State medical marijuana laws, cannabis use and cannabis use disorder among adults with elevated psychological distress. Drug Alcohol Depend 215, 108191. 10.1016/j.drugalcdep.2020.108191.

39. Hughes, J.R., Naud, S., Budney, A.J., Fingar, J.R., and Callas, P.W. (2016). Attempts to stop or reduce daily cannabis use: An intensive natural history study. Psychol Addict Behav 30, 389–397. 10.1037/adb0000155.

40. Mallard, T.T., and Sanchez-Roige, S. (2021). Dimensional Phenotypes in Psychiatric Genetics: Lessons from Genome-Wide Association Studies of Alcohol Use Phenotypes. Complex Psychiatry 7, 45–48. 10.1159/000518863.

41. Fogel, A.I., Akins, M.R., Krupp, A.J., Stagi, M., Stein, V., and Biederer, T. (2007). SynCAMs organize synapses through heterophilic adhesion. J Neurosci 27, 12516–12530. 10.1523/JNEUROSCI.2739-07.2007.

42. Yan, X., Wang, Z., Schmidt, V., Gauert, A., Willnow, T.E., Heinig, M., and Poy, M.N. (2018). Cadm2 regulates body weight and energy homeostasis in mice. Mol Metab 8, 180–188. 10.1016/j.molmet.2017.11.010.

43. Tyler, R.E., Besheer, J., and Joffe, M.E. (2022). Advances in translating mGlu(2) and mGlu(3) receptor selective allosteric modulators as breakthrough treatments for affective disorders and alcohol use disorder. Pharmacol Biochem Behav 219, 173450. 10.1016/j.pbb.2022.173450.

44. Karlsson Linner, R., Mallard, T.T., Barr, P.B., Sanchez-Roige, S., Madole, J.W., Driver, M.N., Poore, H.E., de Vlaming, R., Grotzinger, A.D., Tielbeek, J.J., et al. (2021). Multivariate analysis of 1.5 million people identifies genetic associations with traits related to self-regulation and addiction. Nat Neurosci 24, 1367–1376. 10.1038/s41593-021-00908-3.

45. Agrawal, A., Budney, A.J., and Lynskey, M.T. (2012). The co-occurring use and misuse of cannabis and tobacco: a review. Addiction 107, 1221–1233. 10.1111/j.1360-0443.2012.03837.x.

46. Galimberti, M., Levey, D.F., Deak, J.D., Zhou, H., Stein, M.B., and Gelernter, J. (2024). Genetic influences and causal pathways shared between cannabis use disorder and other substance use traits. Mol Psychiatry. 10.1038/s41380-024-02548-y.

47. Poore, H.E., Hatoum, A., Mallard, T.T., Sanchez-Roige, S., Waldman, I.D., Palmer, A.A., Harden, K.P., Barr, P.B., and Dick, D.M. (2023). A multivariate approach to understanding the genetic overlap between externalizing phenotypes and substance use disorders. Addict Biol 28, e13319. 10.1111/adb.13319.

48. Sanchez-Roige, S., Jennings, M.V., Thorpe, H.H.A., Mallari, J.E., van der Werf, L.C., Bianchi, S.B., Huang, Y., Lee, C., Mallard, T.T., Barnes, S.A., et al. (2023). CADM2 is implicated in impulsive personality and numerous other traits by genome- and phenome-wide association studies in humans and mice. Transl Psychiatry 13, 167. 10.1038/s41398-023-02453-y.

49. Jang, S.K., Saunders, G., Liu, M., and Me Research, T., Jiang, Y., Liu, D.J., and Vrieze, S. (2022). Genetic correlation, pleiotropy, and causal associations between substance use and psychiatric disorder. Psychol Med 52, 968–978. 10.1017/S003329172000272X.

50. Chang, L.H., Ong, J.S., An, J., Verweij, K.J.H., Vink, J.M., Pasman, J., Liu, M., MacGregor, S., Cornelis, M.C., Martin, N.G., and Derks, E.M. (2020). Investigating the genetic and causal relationship between initiation or use of alcohol, caffeine, cannabis and nicotine. Drug Alcohol Depend 210, 107966. 10.1016/j.drugalcdep.2020.107966.

51. Hatoum, A.S., Colbert, S.M.C., Johnson, E.C., Huggett, S.B., Deak, J.D., Pathak, G., Jennings, M.V., Paul, S.E., Karcher, N.R., Hansen, I., et al. (2023). Multivariate genome-wide association meta-analysis of over 1 million subjects identifies loci underlying multiple substance use disorders. Nat Ment Health 1, 210–223. 10.1038/s44220-023-00034-y.

52. Abdellaoui, A., Smit, D.J.A., van den Brink, W., Denys, D., and Verweij, K.J.H. (2021). Genomic relationships across psychiatric disorders including substance use disorders. Drug Alcohol Depend 220, 108535. 10.1016/j.drugalcdep.2021.108535.

53. Waldman, I.D., Poore, H.E., Luningham, J.M., and Yang, J. (2020). Testing structural models of psychopathology at the genomic level. World Psychiatry 19, 350–359. 10.1002/wps.20772.

54. Mallard, T.T., Savage, J.E., Johnson, E.C., Huang, Y., Edwards, A.C., Hottenga, J.J., Grotzinger, A.D., Gustavson, D.E., Jennings, M.V., Anokhin, A., et al. (2022). Item-Level Genome-Wide Association Study of the Alcohol Use Disorders Identification Test in Three Population-Based Cohorts. Am J Psychiatry 179, 58–70. 10.1176/appi.ajp.2020.20091390.

55. Kranzler, H.R., Zhou, H., Kember, R.L., Vickers Smith, R., Justice, A.C., Damrauer, S., Tsao, P.S., Klarin, D., Baras, A., Reid, J., et al. (2019). Genome-wide association study of alcohol consumption and use disorder in 274,424 individuals from multiple populations. Nat Commun 10, 1499. 10.1038/s41467-019-09480-8.

56. Toikumo, S., Jennings, M.V., Pham, B.K., Lee, H., Mallard, T.T., Bianchi, S.B., Meredith, J.J., Vilar-Ribo, L., Xu, H., Hatoum, A.S., et al. (2024). Multi-ancestry meta-analysis of tobacco use disorder identifies 461 potential risk genes and reveals associations with multiple health outcomes. Nature Human Behavior. 10.1038/s41562-024-01851-6.

57. Zhou, H., Kember, R.L., Deak, J.D., Xu, H., Toikumo, S., Yuan, K., Lind, P.A., Farajzadeh, L., Wang, L., Hatoum, A.S., et al. (2023). Multi-ancestry study of the genetics of problematic alcohol use in over 1 million individuals. Nat Med 29, 3184–3192. 10.1038/s41591-023-02653-5.

58. Sanchez-Roige, S., Palmer, A.A., Fontanillas, P., Elson, S.L., 23andMe Research Team, Substance Use Disorder Working Group of the Psychiatric Genomics Consortium, Adams, M.J., Howard, D.M., Edenberg, H.J., Davies, G., et al. (2019). Genome-Wide Association Study Meta-Analysis of the Alcohol Use Disorders Identification Test (AUDIT) in Two Population-Based Cohorts. Am J Psychiatry 176, 107–118. 10.1176/appi.ajp.2018.18040369.

59. Savage, J.E., Barr, P.B., Phung, T., Lee, Y.H., Zhang, Y., Ge, T., Smoller, J.W., Davis, L.K., Meyers, J., Porjesz, B., et al. (2023). Genetic Heterogeneity Across Dimensions of Alcohol Use Behaviors. medRxiv, 2023.2012.2026.23300537. 10.1101/2023.12.26.23300537.

60. Saunders, G.R.B., Wang, X., Chen, F., Jang, S.K., Liu, M., Wang, C., Gao, S., Jiang, Y., Khunsriraksakul, C., Otto, J.M., et al. (2022). Genetic diversity fuels gene discovery for tobacco and alcohol use. Nature 612, 720–724. 10.1038/s41586-022-05477-4.

61. Trubetskoy, V., Pardinas, A.F., Qi, T., Panagiotaropoulou, G., Awasthi, S., Bigdeli, T.B., Bryois, J., Chen, C.Y., Dennison, C.A., Hall, L.S., et al. (2022). Mapping genomic loci implicates genes and synaptic biology in schizophrenia. Nature 604, 502–508. 10.1038/s41586-022-04434-5.

62. Lam, M., Chen, C.Y., Li, Z., Martin, A.R., Bryois, J., Ma, X., Gaspar, H., Ikeda, M., Benyamin, B., Brown, B.C., et al. (2019). Comparative genetic architectures of schizophrenia in East Asian and European populations. Nat Genet 51, 1670–1678. 10.1038/s41588-019-0512-x.

63. Pardinas, A.F., Holmans, P., Pocklington, A.J., Escott-Price, V., Ripke, S., Carrera, N., Legge, S.E., Bishop, S., Cameron, D., Hamshere, M.L., et al. (2018). Common schizophrenia alleles are enriched in mutation-intolerant genes and in regions under strong background selection. Nat Genet 50, 381–389. 10.1038/s41588-018-0059-2.

64. Li, Z., Chen, J., Yu, H., He, L., Xu, Y., Zhang, D., Yi, Q., Li, C., Li, X., Shen, J., et al. (2017). Genome-wide association analysis identifies 30 new susceptibility loci for schizophrenia. Nat Genet 49, 1576–1583. 10.1038/ng.3973.

65. Schizophrenia Working Group of the Psychiatric Genomics Consortium (2014). Biological insights from 108 schizophrenia-associated genetic loci. Nature 511, 421–427. 10.1038/nature13595.

66. Nagel, M., Watanabe, K., Stringer, S., Posthuma, D., and van der Sluis, S. (2018). Item-level analyses reveal genetic heterogeneity in neuroticism. Nat Commun 9, 905. 10.1038/s41467-018-03242-8.

67. Luciano, M., Hagenaars, S.P., Davies, G., Hill, W.D., Clarke, T.K., Shirali, M., Harris, S.E., Marioni, R.E., Liewald, D.C., Fawns-Ritchie, C., et al. (2018). Association analysis in over 329,000 individuals identifies 116 independent variants influencing neuroticism. Nat Genet 50, 6–11. 10.1038/s41588-017-0013-8.

68. Lee, J.J., Wedow, R., Okbay, A., Kong, E., Maghzian, O., Zacher, M., Nguyen-Viet, T.A., Bowers, P., Sidorenko, J., Karlsson Linner, R., et al. (2018). Gene discovery and polygenic prediction from a genome-wide association study of educational attainment in 1.1 million individuals. Nat Genet 50, 1112–1121. 10.1038/s41588-018-0147-3.

69. Hysi, P.G., Choquet, H., Khawaja, A.P., Wojciechowski, R., Tedja, M.S., Yin, J., Simcoe, M.J., Patasova, K., Mahroo, O.A., Thai, K.K., et al. (2020). Meta-analysis of 542,934 subjects of European ancestry identifies new genes and mechanisms predisposing to refractive error and myopia. Nat Genet 52, 401–407. 10.1038/s41588-020-0599-0.

70. Tedja, M.S., Wojciechowski, R., Hysi, P.G., Eriksson, N., Furlotte, N.A., Verhoeven, V.J.M., Iglesias, A.I., Meester-Smoor, M.A., Tompson, S.W., Fan, Q., et al. (2018). Genome-wide association meta-analysis highlights light-induced signaling as a driver for refractive error. Nat Genet 50, 834–848. 10.1038/s41588-018-0127-7.

71. Watanabe, K., Jansen, P.R., Savage, J.E., Nandakumar, P., Wang, X., 23andMe Research Team, Hinds, D.A., Gelernter, J., Levey, D.F., Polimanti, R., et al. (2022). Genome-wide meta-analysis of insomnia prioritizes genes associated with metabolic and psychiatric pathways. Nat Genet 54, 1125–1132. 10.1038/s41588-022-01124-w.

72. Di Menna, L., Joffe, M.E., Iacovelli, L., Orlando, R., Lindsley, C.W., Mairesse, J., Gressens, P., Cannella, M., Caraci, F., Copani, A., et al. (2018). Functional partnership between mGlu3 and mGlu5 metabotropic glutamate receptors in the central nervous system. Neuropharmacology 128, 301–313. 10.1016/j.neuropharm.2017.10.026.

73. Ibrahim, K.S., Abd-Elrahman, K.S., El Mestikawy, S., and Ferguson, S.S.G. (2020). Targeting Vesicular Glutamate Transporter Machinery: Implications on Metabotropic Glutamate Receptor 5 Signaling and Behavior. Mol Pharmacol 98, 314–327. 10.1124/molpharm.120.000089.

74. Jung, K.M., Mangieri, R., Stapleton, C., Kim, J., Fegley, D., Wallace, M., Mackie, K., and Piomelli, D. (2005). Stimulation of endocannabinoid formation in brain slice cultures through activation of group I metabotropic glutamate receptors. Mol Pharmacol 68, 1196–1202. 10.1124/mol.105.013961.

75. The GTEx Consortium (2024). GTEx Portal. https://gtexportal.org/.

76. Schoeler, T., Speed, D., Porcu, E., Pirastu, N., Pingault, J.B., and Kutalik, Z. (2023). Participation bias in the UK Biobank distorts genetic associations and downstream analyses. Nat Hum Behav 7, 1216–1227. 10.1038/s41562-023-01579-9.

77. Deak, J.D., Levey, D.F., Wendt, F.R., Zhou, H., Galimberti, M., Kranzler, H.R., Gaziano, J.M., Stein, M.B., Polimanti, R., Gelernter, J., and Million Veteran, P. (2022). Genome-Wide Investigation of Maximum Habitual Alcohol Intake in US Veterans in Relation to Alcohol Consumption Traits and Alcohol Use Disorder. JAMA Netw Open 5, e2238880. 10.1001/jamanetworkopen.2022.38880.

78. Pasman, J.A., Demange, P.A., Guloksuz, S., Willemsen, A.H.M., Abdellaoui, A., Ten Have, M., Hottenga, J.J., Boomsma, D.I., de Geus, E., Bartels, M., et al. (2022). Genetic Risk for Smoking: Disentangling Interplay Between Genes and Socioeconomic Status. Behav Genet 52, 92–107. 10.1007/s10519-021-10094-4.

79. Xu, K., Li, B., McGinnis, K.A., Vickers-Smith, R., Dao, C., Sun, N., Kember, R.L., Zhou, H., Becker, W.C., Gelernter, J., et al. (2020). Genome-wide association study of smoking trajectory and meta-analysis of smoking status in 842,000 individuals. Nat Commun 11, 5302. 10.1038/s41467-020-18489-3.

80. Zhou, H., Sealock, J.M., Sanchez-Roige, S., Clarke, T.K., Levey, D.F., Cheng, Z., Li, B., Polimanti, R., Kember, R.L., Smith, R.V., et al. (2020). Genome-wide meta-analysis of problematic alcohol use in 435,563 individuals yields insights into biology and relationships with other traits. Nat Neurosci 23, 809–818. 10.1038/s41593-020-0643-5.

81. Cai, N., Revez, J.A., Adams, M.J., Andlauer, T.F.M., Breen, G., Byrne, E.M., Clarke, T.K., Forstner, A.J., Grabe, H.J., Hamilton, S.P., et al. (2020). Minimal phenotyping yields genome-wide association signals of low specificity for major depression. Nat Genet 52, 437–447. 10.1038/s41588-020-0594-5.

82. Evangelou, E., Gao, H., Chu, C., Ntritsos, G., Blakeley, P., Butts, A.R., Pazoki, R., Suzuki, H., Koskeridis, F., Yiorkas, A.M., et al. (2019). New alcohol-related genes suggest shared genetic mechanisms with neuropsychiatric disorders. Nat Hum Behav 3, 950–961. 10.1038/s41562-019-0653-z.

83. Zhong, V.W., Kuang, A., Danning, R.D., Kraft, P., van Dam, R.M., Chasman, D.I., and Cornelis, M.C. (2019). A genome-wide association study of bitter and sweet beverage consumption. Hum Mol Genet 28, 2449–2457. 10.1093/hmg/ddz061.

84. Sanchez-Roige, S., Fontanillas, P., Elson, S.L., Gray, J.C., de Wit, H., MacKillop, J., and Palmer, A.A. (2019). Genome-Wide Association Studies of Impulsive Personality Traits (BIS-11 and UPPS-P) and Drug Experimentation in up to 22,861 Adult Research Participants Identify Loci in the CACNA1I and CADM2 genes. J Neurosci 39, 2562–2572. 10.1523/JNEUROSCI.2662-18.2019.

85. Karlsson Linner, R., Biroli, P., Kong, E., Meddens, S.F.W., Wedow, R., Fontana, M.A., Lebreton, M., Tino, S.P., Abdellaoui, A., Hammerschlag, A.R., et al. (2019). Genome-wide association analyses of risk tolerance and risky behaviors in over 1 million individuals identify hundreds of loci and shared genetic influences. Nat Genet 51, 245–257. 10.1038/s41588-018-0309-3.

86. Liu, M., Jiang, Y., Wedow, R., Li, Y., Brazel, D.M., Chen, F., Datta, G., Davila-Velderrain, J., McGuire, D., Tian, C., et al. (2019). Association studies of up to 1.2 million individuals yield new insights into the genetic etiology of tobacco and alcohol use. Nat Genet 51, 237–244. 10.1038/s41588-018-0307-5.

87. Erzurumluoglu, A.M., Liu, M., Jackson, V.E., Barnes, D.R., Datta, G., Melbourne, C.A., Young, R., Batini, C., Surendran, P., Jiang, T., et al. (2020). Meta-analysis of up to 622,409 individuals identifies 40 novel smoking behaviour associated genetic loci. Mol Psychiatry 25, 2392–2409. 10.1038/s41380-018-0313-0.

88. Kichaev, G., Bhatia, G., Loh, P.R., Gazal, S., Burch, K., Freund, M.K., Schoech, A., Pasaniuc, B., and Price, A.L. (2019). Leveraging Polygenic Functional Enrichment to Improve GWAS Power. Am J Hum Genet 104, 65–75. 10.1016/j.ajhg.2018.11.008.

89. Clifton, E.A.D., Perry, J.R.B., Imamura, F., Lotta, L.A., Brage, S., Forouhi, N.G., Griffin, S.J., Wareham, N.J., Ong, K.K., and Day, F.R. (2018). Genome-wide association study for risk taking propensity indicates shared pathways with body mass index. Commun Biol 1, 36. 10.1038/s42003-018-0042-6.

90. Clarke, T.K., Adams, M.J., Davies, G., Howard, D.M., Hall, L.S., Padmanabhan, S., Murray, A.D., Smith, B.H., Campbell, A., Hayward, C., et al. (2017). Genome-wide association study of alcohol consumption and genetic overlap with other health-related traits in UK Biobank (N=112 117). Mol Psychiatry 22, 1376–1384. 10.1038/mp.2017.153.

91. Brazel, D.M., Jiang, Y., Hughey, J.M., Turcot, V., Zhan, X., Gong, J., Batini, C., Weissenkampen, J.D., Liu, M., CHD Exome+ Consortium Consortium for Genetics of Smoking Behaviour, et al. (2019). Exome Chip Meta-analysis Fine Maps Causal Variants and Elucidates the Genetic Architecture of Rare Coding Variants in Smoking and Alcohol Use. Biol Psychiatry 85, 946–955. 10.1016/j.biopsych.2018.11.024.

92. Baselmans, B., Hammerschlag, A.R., Noordijk, S., Ip, H., van der Zee, M., de Geus, E., Abdellaoui, A., Treur, J.L., and van ’t Ent, D. (2022). The Genetic and Neural Substrates of Externalizing Behavior. Biol Psychiatry Glob Open Sci 2, 389–399. 10.1016/j.bpsgos.2021.09.007.

93. Strawbridge, R.J., Ward, J., Cullen, B., Tunbridge, E.M., Hartz, S., Bierut, L., Horton, A., Bailey, M.E.S., Graham, N., Ferguson, A., et al. (2018). Genome-wide analysis of self-reported risk-taking behaviour and cross-disorder genetic correlations in the UK Biobank cohort. Transl Psychiatry 8, 39. 10.1038/s41398-017-0079-1.

94. Mills, M.C., Tropf, F.C., Brazel, D.M., van Zuydam, N., Vaez, A., eQTLGen Consortium, Bios Consortium, Human Reproductive Behaviour Consortium, Pers, T.H., Snieder, H., et al. (2021). Identification of 371 genetic variants for age at first sex and birth linked to externalising behaviour. Nat Hum Behav 5, 1717–1730. 10.1038/s41562-021-01135-3.

95. Johnson, E.C., Sanchez-Roige, S., Acion, L., Adams, M.J., Bucholz, K.K., Chan, G., Chao, M.J., Chorlian, D.B., Dick, D.M., Edenberg, H.J., et al. (2021). Polygenic contributions to alcohol use and alcohol use disorders across population-based and clinically ascertained samples. Psychol Med 51, 1147–1156. 10.1017/S0033291719004045.

96. Sun, Y., Chang, S., Wang, F., Sun, H., Ni, Z., Yue, W., Zhou, H., Gelernter, J., Malison, R.T., Kalayasiri, R., et al. (2019). Genome-wide association study of alcohol dependence in male Han Chinese and cross-ethnic polygenic risk score comparison. Transl Psychiatry 9, 249. 10.1038/s41398-019-0586-3.

97. Lai, D., Wetherill, L., Bertelsen, S., Carey, C.E., Kamarajan, C., Kapoor, M., Meyers, J.L., Anokhin, A.P., Bennett, D.A., Bucholz, K.K., et al. (2019). Genome-wide association studies of alcohol dependence, DSM-IV criterion count and individual criteria. Genes Brain Behav 18, e12579. 10.1111/gbb.12579.

98. Chang, L.H., Couvy-Duchesne, B., Liu, M., Medland, S.E., Verhulst, B., Benotsch, E.G., Hickie, I.B., Martin, N.G., Gillespie, N.A., and GSCAN Consortium (2019). Association between polygenic risk for tobacco or alcohol consumption and liability to licit and illicit substance use in young Australian adults. Drug Alcohol Depend 197, 271–279. 10.1016/j.drugalcdep.2019.01.015.

99. Allegrini, A.G., Verweij, K.J.H., Abdellaoui, A., Treur, J.L., Hottenga, J.J., Willemsen, G., Boomsma, D.I., International Cannabis Consortium, and Vink, J.M. (2019). Genetic Vulnerability for Smoking and Cannabis Use: Associations With E-Cigarette and Water Pipe Use. Nicotine Tob Res 21, 723–730. 10.1093/ntr/nty150.

100. Hodgson, K., Coleman, J.R.I., Hagenaars, S.P., Purves, K.L., Glanville, K., Choi, S.W., O’Reilly, P., Breen, G., Major Depressive Disorder Working Group of the Psychiatric Genomics Consortium, and Lewis, C.M. (2020). Cannabis use, depression and self-harm: phenotypic and genetic relationships. Addiction 115, 482–492. 10.1111/add.14845.

101. Smeland, O.B., and Andreassen, O.A. (2021). Polygenic risk scores in psychiatry - Large potential but still limited clinical utility. Eur Neuropsychopharmacol 51, 68–70. 10.1016/j.euroneuro.2021.05.007.

102. Kember, R.L., Hartwell, E.E., Xu, H., Rotenberg, J., Almasy, L., Zhou, H., Gelernter, J., and Kranzler, H.R. (2023). Phenome-wide Association Analysis of Substance Use Disorders in a Deeply Phenotyped Sample. Biol Psychiatry 93, 536–545. 10.1016/j.biopsych.2022.08.010.

103. Cheng, W., Parker, N., Karadag, N., Koch, E., Hindley, G., Icick, R., Shadrin, A., O’Connell, K.S., Bjella, T., Bahrami, S., et al. (2023). The relationship between cannabis use, schizophrenia, and bipolar disorder: a genetically informed study. Lancet Psychiatry 10, 441–451. 10.1016/S2215-0366(23)00143-8.

104. Johnson, E.C., Hatoum, A.S., Deak, J.D., Polimanti, R., Murray, R.M., Edenberg, H.J., Gelernter, J., Di Forti, M., and Agrawal, A. (2021). The relationship between cannabis and schizophrenia: a genetically informed perspective. Addiction 116, 3227–3234. 10.1111/add.15534.

105. Reginsson, G.W., Ingason, A., Euesden, J., Bjornsdottir, G., Olafsson, S., Sigurdsson, E., Oskarsson, H., Tyrfingsson, T., Runarsdottir, V., Hansdottir, I., et al. (2018). Polygenic risk scores for schizophrenia and bipolar disorder associate with addiction. Addict Biol 23, 485–492. 10.1111/adb.12496.

106. Johnson, E.C., Austin-Zimmerman, I., Thorpe, H.H.A., Levey, D.F., Baranger, D.A.A., Colbert, S.M.C., Thorgeirsson, T., Khokhar, J.Y., Davis, L.K., Di Forti, M., et al. (2024). Dissecting the relationships between tobacco smoking, cannabis use disorder, and schizophrenia using genomic approaches.

107. Hamilton, I., and Monaghan, M. (2019). Cannabis and Psychosis: Are We any Closer to Understanding the Relationship? Curr Psychiatry Rep 21, 48. 10.1007/s11920-019-1044-x.

108. Di Forti, M., Quattrone, D., Freeman, T.P., Tripoli, G., Gayer-Anderson, C., Quigley, H., Rodriguez, V., Jongsma, H.E., Ferraro, L., La Cascia, C., et al. (2019). The contribution of cannabis use to variation in the incidence of psychotic disorder across Europe (EU-GEI): a multicentre case-control study. Lancet Psychiatry 6, 427–436. 10.1016/S2215-0366(19)30048-3.

109. Oluwoye, O., Monroe-DeVita, M., Burduli, E., Chwastiak, L., McPherson, S., McClellan, J.M., and McDonell, M.G. (2019). Impact of tobacco, alcohol and cannabis use on treatment outcomes among patients experiencing first episode psychosis: Data from the national RAISE-ETP study. Early Interv Psychiatry 13, 142–146. 10.1111/eip.12542.

110. Schoeler, T., Monk, A., Sami, M.B., Klamerus, E., Foglia, E., Brown, R., Camuri, G., Altamura, A.C., Murray, R., and Bhattacharyya, S. (2016). Continued versus discontinued cannabis use in patients with psychosis: a systematic review and meta-analysis. Lancet Psychiatry 3, 215–225. 10.1016/S2215-0366(15)00363-6.

111. Durvasula, A., and Price, A.L. (2023). Distinct explanations underlie gene-environment interactions in the UK Biobank. medRxiv. 10.1101/2023.09.22.23295969.

112. Abdellaoui, A., Dolan, C.V., Verweij, K.J.H., and Nivard, M.G. (2022). Gene-environment correlations across geographic regions affect genome-wide association studies. Nat Genet 54, 1345–1354. 10.1038/s41588-022-01158-0.

113. Thorpe, H.H.A., Fontanillas, P., Pham, B.K., Meredith, J.J., Jennings, M.V., Courchesne-Krak, N.S., Vilar-Ribó, L., Bianchi, S.B., Mutz, J., 23andMe Research Team, et al. (2024). Genome-wide association studies of coffee intake in UK/US participants of European ancestry uncover cohort-specific genetic associations. Neuropsychopharmacology. 10.1038/s41386-024-01870-x.

114. Kayir, H., Ruffolo, J., McCunn, P., and Khokhar, J.Y. (2023). The Relationship Between Cannabis, Cognition, and Schizophrenia: It’s Complicated. Curr Top Behav Neurosci 63, 437–461. 10.1007/7854_2022_396.

115. Farrelly, K.N., Wardell, J.D., Marsden, E., Scarfe, M.L., Najdzionek, P., Turna, J., and MacKillop, J. (2023). The Impact of Recreational Cannabis Legalization on Cannabis Use and Associated Outcomes: A Systematic Review. Subst Abuse 17, 11782218231172054. 10.1177/11782218231172054.

116. Fergusson, D.M., Boden, J.M., and Horwood, L.J. (2015). Psychosocial sequelae of cannabis use and implications for policy: findings from the Christchurch Health and Development Study. Soc Psychiatry Psychiatr Epidemiol 50, 1317–1326. 10.1007/s00127-015-1070-x.

117. van der Pol, P., Liebregts, N., de Graaf, R., Korf, D.J., van den Brink, W., and van Laar, M. (2013). Predicting the transition from frequent cannabis use to cannabis dependence: a three-year prospective study. Drug Alcohol Depend 133, 352–359. 10.1016/j.drugalcdep.2013.06.009.

118. Walden, N., and Earleywine, M. (2008). How high: quantity as a predictor of cannabis-related problems. Harm Reduct J 5, 20. 10.1186/1477-7517-5-20.

119. Zeisser, C., Thompson, K., Stockwell, T., Duff, C., Chow, C., Vallance, K., Ivsins, A., Michelow, W., Marsh, D., and Lucas, P. (2012). A ‘standard joint’? The role of quantity in predicting cannabis-related problems. Addiction Research & Theory 20, 82–92. 10.3109/16066359.2011.569101.

120. Substance Abuse and Mental Health Services Administration (2022). Key substance use and mental health indicators in the United States: Results from the 2022 National Survey on Drug Use and Health. Center for Behavioral Health Statistics and Quality, Substance Abuse and Mental Health Services Administration. https://www.samhsa.gov/data/sites/default/files/reports/rpt42731/2022-nsduh-nnr.pdf.

121. Althubaiti, A. (2016). Information bias in health research: definition, pitfalls, and adjustment methods. J Multidiscip Healthc 9, 211–217. 10.2147/JMDH.S104807.

122. Jeffers, A.M., Glantz, S., Byers, A., and Keyhani, S. (2021). Sociodemographic Characteristics Associated With and Prevalence and Frequency of Cannabis Use Among Adults in the US. JAMA Netw Open 4, e2136571. 10.1001/jamanetworkopen.2021.36571.

123. Karriker-Jaffe, K.J. (2013). Neighborhood socioeconomic status and substance use by U.S. adults. Drug Alcohol Depend 133, 212–221. 10.1016/j.drugalcdep.2013.04.033.

124. Atkinson, E.G., Bianchi, S.B., Ye, G.Y., Martinez-Magana, J.J., Tietz, G.E., Montalvo-Ortiz, J.L., Giusti-Rodriguez, P., Palmer, A.A., and Sanchez-Roige, S. (2022). Cross-ancestry genomic research: time to close the gap. Neuropsychopharmacology 47, 1737–1738. 10.1038/s41386-022-01365-7.

125. Martin, A.R., Kanai, M., Kamatani, Y., Okada, Y., Neale, B.M., and Daly, M.J. (2019). Clinical use of current polygenic risk scores may exacerbate health disparities. Nat Genet 51, 584–591. 10.1038/s41588-019-0379-x.

126. Bryc, K., Durand, E.Y., Macpherson, J.M., Reich, D., and Mountain, J.L. (2015). The genetic ancestry of African Americans, Latinos, and European Americans across the United States. Am J Hum Genet 96, 37–53. 10.1016/j.ajhg.2014.11.010.

127. National Academies of Sciences, Engineering, and Medicine Health and Medicine Division, Division of Behavioral and Social Sciences and Education, Board on Health Sciences Policy, Committee on Population, Committee on the Use of Race, and Ethnicity, and Ancestry as Population Descriptors in Genomics Research (2023). In Using Population Descriptors in Genetics and Genomics Research: A New Framework for an Evolving Field. 10.17226/26902.

128. Durand, E.Y., Do, C.B., Mountain, J.L., and Macpherson, J.M. (2014). Ancestry Composition: A Novel, Efficient Pipeline for Ancestry Deconvolution. bioRxiv. [Preprint]. 10.1101/010512.

129. Yin, B., Wang, X., Huang, T., and Jia, J. (2022). Shared Genetics and Causality Between Decaffeinated Coffee Consumption and Neuropsychiatric Diseases: A Large-Scale Genome-Wide Cross-Trait Analysis and Mendelian Randomization Analysis. Front Psychiatry 13, 910432. 10.3389/fpsyt.2022.910432.

130. Agrawal, A., Chou, Y.L., Carey, C.E., Baranger, D.A.A., Zhang, B., Sherva, R., Wetherill, L., Kapoor, M., Wang, J.C., Bertelsen, S., et al. (2018). Genome-wide association study identifies a novel locus for cannabis dependence. Mol Psychiatry 23, 1293–1302. 10.1038/mp.2017.200.

131. Sherva, R., Wang, Q., Kranzler, H., Zhao, H., Koesterer, R., Herman, A., Farrer, L.A., and Gelernter, J. (2016). Genome-wide Association Study of Cannabis Dependence Severity, Novel Risk Variants, and Shared Genetic Risks. JAMA Psychiatry 73, 472–480. 10.1001/jamapsychiatry.2016.0036.

132. Agrawal, A., Lynskey, M.T., Hinrichs, A., Grucza, R., Saccone, S.F., Krueger, R., Neuman, R., Howells, W., Fisher, S., Fox, L., et al. (2011). A genome-wide association study of DSM-IV cannabis dependence. Addict Biol 16, 514–518. 10.1111/j.1369-1600.2010.00255.x.

133. Minica, C.C., Dolan, C.V., Hottenga, J.J., Pool, R., Genome of the Netherlands Consortium, Fedko, I.O., Mbarek, H., Huppertz, C., Bartels, M., Boomsma, D.I., and Vink, J.M. (2015). Heritability, SNP- and Gene-Based Analyses of Cannabis Use Initiation and Age at Onset. Behav Genet 45, 503–513. 10.1007/s10519-015-9723-9.

134. Zhao, Y., Han, X., and Zheng, Z.L. (2023). Analysis of the brain transcriptome for substance-associated genes: An update on large-scale genome-wide association studies. Addict Biol 28, e13332. 10.1111/adb.13332.

135. Greco, L.A., Reay, W.R., Dayas, C.V., and Cairns, M.J. (2023). Exploring opportunities for drug repurposing and precision medicine in cannabis use disorder using genetics. Addict Biol 28, e13313. 10.1111/adb.13313.

136. Carreras-Gallo, N., Dwaraka, V.B., Caceres, A., Smith, R., Mendez, T.L., Went, H., and Gonzalez, J.R. (2023). Impact of tobacco, alcohol, and marijuana on genome-wide DNA methylation and its relationship with hypertension. Epigenetics 18, 2214392. 10.1080/15592294.2023.2214392.

137. Greco, L.A., Reay, W.R., Dayas, C.V., and Cairns, M.J. (2022). Pairwise genetic meta-analyses between schizophrenia and substance dependence phenotypes reveals novel association signals with pharmacological significance. Transl Psychiatry 12, 403. 10.1038/s41398-022-02186-4.

138. Minica, C.C., Verweij, K.J.H., van der Most, P.J., Mbarek, H., Bernard, M., van Eijk, K.R., Lind, P.A., Liu, M.Z., Maciejewski, D.F., Palviainen, T., et al. (2018). Genome-wide association meta-analysis of age at first cannabis use. Addiction 113, 2073–2086. 10.1111/add.14368.

139. Sey, N.Y.A., Hu, B., Mah, W., Fauni, H., McAfee, J.C., Rajarajan, P., Brennand, K.J., Akbarian, S., and Won, H. (2020). A computational tool (H-MAGMA) for improved prediction of brain-disorder risk genes by incorporating brain chromatin interaction profiles. Nat Neurosci 23, 583–593. 10.1038/s41593-020-0603-0.

140. Barbeira, A.N., Dickinson, S.P., Bonazzola, R., Zheng, J., Wheeler, H.E., Torres, J.M., Torstenson, E.S., Shah, K.P., Garcia, T., Edwards, T.L., et al. (2018). Exploring the phenotypic consequences of tissue specific gene expression variation inferred from GWAS summary statistics. Nat Commun 9, 1825. 10.1038/s41467-018-03621-1.

141. Bulik-Sullivan, B.K., Loh, P.R., Finucane, H.K., Ripke, S., Yang, J., Schizophrenia Working Group of the Psychiatric Genomics Consortium, Patterson, N., Daly, M.J., Price, A.L., and Neale, B.M. (2015). LD Score regression distinguishes confounding from polygenicity in genome-wide association studies. Nat Genet 47, 291–295. 10.1038/ng.3211.

142. All of Us Research Program Genomics Investigators (2024). Genomic data in the All of Us Research Program. Nature 627, 340–346. 10.1038/s41586-023-06957-x.

143. All of Us Research Program Investigators, Denny, J.C., Rutter, J.L., Goldstein, D.B., Philippakis, A., Smoller, J.W., Jenkins, G., and Dishman, E. (2019). The “All of Us” Research Program. N Engl J Med 381, 668–676. 10.1056/NEJMsr1809937.

144. Ge, T., Chen, C.Y., Ni, Y., Feng, Y.A., and Smoller, J.W. (2019). Polygenic prediction via Bayesian regression and continuous shrinkage priors. Nat Commun 10, 1776. 10.1038/s41467-019-09718-5.

145. Lee, S.H., Goddard, M.E., Wray, N.R., and Visscher, P.M. (2012). A better coefficient of determination for genetic profile analysis. Genet Epidemiol 36, 214–224. 10.1002/gepi.21614.

146. Roden, D.M., Pulley, J.M., Basford, M.A., Bernard, G.R., Clayton, E.W., Balser, J.R., and Masys, D.R. (2008). Development of a large-scale de-identified DNA biobank to enable personalized medicine. Clin Pharmacol Ther 84, 362–369. 10.1038/clpt.2008.89.

147. Dennis, J., Sealock, J., Levinson, R.T., Farber-Eger, E., Franco, J., Fong, S., Straub, P., Hucks, D., Song, W.L., Linton, M.F., et al. (2021). Genetic risk for major depressive disorder and loneliness in sex-specific associations with coronary artery disease. Mol Psychiatry 26, 4254–4264. 10.1038/s41380-019-0614-y.

