## Supplementary Material for "Genome-wide association studies of lifetime and frequency cannabis use in 131,895 individuals"

### Supplementary Methods

#### Analysis of local ancestry

We performed lifetime cannabis use GWAS (*N*=131,895) and frequency of cannabis use GWAS (*N*=73,374) on 23andMe, Inc. participants classified as being of European ancestry. Ancestry falls along a spectrum[1,2]; each individual was clustered based on genetic similarity to a reference panel using local ancestry analysis[3]. Briefly, the 23andMe algorithm first partitions phased genomic data into short windows of about 300 SNPs. Within each window, we use a support vector machine to classify individual haplotypes into one of 31 reference populations (https://www.23andme.com/ancestry-composition-guide/). The support vector machine classifications are fed into a hidden Markov model that accounts for switch errors and incorrect assignments, and gives probabilities for each reference population in each window. The reference population data is derived from public datasets (the Human Genome Diversity Project, HapMap, and 1000 Genomes), as well as 23andMe customers who have reported having four grandparents from the same country.

Ancestries are defined as follows:

| Ancestry | Classification Criteria |
|---|---|
| European | European + Middle Eastern > 0.97, European > 0.90 |
| East Asian | East Asian + Southeast Asian > 0.97 |
| South Asian | South Asian > 0.97 |
| Middle Eastern (& North African) | Middle Eastern + European > 0.97, Middle Eastern > 0.90 |
| African American + Latin American | European + African + East Asian + Native American + Middle Eastern > 0.90, African + Native American > 0.01 |

**Genome-wide association and secondary analyses**

DNA extraction and genotyping were performed from saliva samples by clinical laboratories CLIA-certified and CAP-accredited by the Laboratory Corporation of America. 23andMe, Inc. conducted all quality control, imputation, and univariate genome-wide analyses as previously described (see **Supplementary Table 3** for SNPs analyzed following quality control and imputation)[4,5]. Participants were genotyped on one of five Illumina genotyping platforms, containing between 550,000 to 950,000 variants, for a total of 1.6 million genotyped variants. Samples were genotyped on one of five genotyping platforms. The V1 and V2 platforms were variants of the Illumina HumanHap550 + BeadChip, including about 25,000 custom SNPs selected by 23andMe, with a total of ~560,000 SNPs. The V3 platform was based on the Illumina OmniExpress + BeadChip, with custom content to improve the overlap with our V2 array, with a total of ~950,000 SNPs. The V4 platform is a fully custom array, including a lower redundancy subset of V2 and V3 SNPs with additional coverage of lower-frequency coding variation, and ~570,000 SNPs. The v5 platform, in current use, is an Illumina Infinium Global Screening Array (~640,000 variants) supplemented with ~50,000 variants of custom content. Samples that failed to reach 98.5% call rate were excluded from the study. Samples that failed to reach 98.5% call rate were re-analyzed[6,7]. Variants were imputed based on an imputation panel combining 1000 Genomes Phase 3, UK10K and the Human Reference Consortium (**HRC**). About 64.4 million variants were then imputed against the HRC panel, augmented by a single unified imputation reference panel combining the May 2015 release of the 1000 Genomes Phase 3 haplotypes with the UK10K imputation reference panel for variants not present in the HRC. Imputed variants with low imputation quality ($r^2$<0.50 averaged across batches or a minimum $r^2$<0.30), or with evidence of batch effects ($p$<1.00E-50) were removed[6,7]. A total of 1.3 million genotyped and 30.5 million imputed variants passed the pre- and post-GWAS quality controls. We furthermore filtered out variants with minor allele frequency (**MAF**) <0.1%, which are extremely sensitive to quantitative trait over-dispersion, reducing to 14.1 million variants available for follow-up analyses. Principal components were computed using ~65,000 high-quality genotyped variants present in all five genotyping platforms.

A maximal set of unrelated individuals was chosen for the analysis using a segmental identity-by-descent (**IBD**) estimation algorithm[8] to ensure that only unrelated individuals were included in the sample. Individuals were defined as related if they shared more than 700 cM IBD, including regions where the two individuals shared either one or both genomic segments IBD. This level of relatedness (~20% of the genome) corresponds to approximately the minimal expected sharing between first cousins in an outbred population. For the lifetime cannabis use binary phenotype, if a case was found to be related to a control, the case was preferentially kept in the sample. The 23andMe GWAS pipeline performs linear regression and assumes an additive model for allelic effects[9-12]. Age (inverse-normal transformed), sex, the top five principal genotype components, and indicator variables for genotype platforms were applied as covariates and *p*-values were corrected for genomic control. We imputed participant genotype data against the September 2013 release of 1000 Genomes phase 1 version 3 reference haplotypes. We phased and imputed data for each genotyping platform separately. We phased using an internally developed phasing tool, Finch, which implements the Beagle haplotype graph-based phasing algorithm, modified to separate the haplotype graph construction and phasing steps. In preparation for imputation, we split phased chromosomes into segments of no more than 10,000 genotyped SNPs, with overlaps of 200 SNPs. We excluded SNPs of low genotyping quality, including those that failed a Mendelian transmission test in trios ($p$<1.00E-20) or with large allele frequency discrepancies compared to European 1000 Genomes reference data, failed Hardy-Weinberg test ($p$<1.00E-20), failed batch effects test (ANOVA $p$<1.00E-20), or had a call rate <90%. Frequency discrepancies were identified by computing a 2 x 2 table of allele counts for European 1000 Genomes samples and 2000 randomly sampled 23andMe research participants with European ancestries, and identifying SNPs with a $\chi^2$ $p$<10E-15. We imputed each phased segment against all-ethnicity 1000 Genomes haplotypes (excluding monomorphic and singleton sites) using Minimac2[13], using 5 rounds and 200 states for parameter estimation.

For the X chromosome, we built separate haplotype graphs for the non-pseudoautosomal region and each pseudoautosomal region, and these regions were phased separately. We then imputed males and females together using Minimac2, as with the autosomes, treating males as homozygous pseudo-diploids for the non-pseudoautosomal region.

For tests using imputed data, we use the imputed dosages rather than best-guess genotypes. We imputed HLA allele dosages from SNP genotype data using HIBAG. We imputed alleles for HLA-A, B, C, DPB1, DQA1, DQB1, and DRB1 loci at four-digit resolution. To test associations between HLA allele dosages and phenotypes, we performed linear regression using the same set of covariates used in the SNP based GWAS. We performed separate association tests for each imputed allele. HLA analysis did not identify any significant signal and hence are not described in the main text.

**Genetic correlation, PheWAS, and LabWAS traits**

Genetic correlations were conducted using a set of 292 summary statistics across 22 health, psychiatric, and anthropomorphic categories. Summary statistics related to health included 21 cancer, 16 cardiovascular, 10 immune & inflammation, 10 metabolic, 9 gastrointestinal, 8 neurological, 6 pain, and 15 other health traits. Summary statistics related to psychiatric health included 39 substance use & misuse, 23 anxiety & stress disorder, 23 bipolar & depression, 12 cognitive & executive function, 8 psychosis, 4 suicide, and 17 other psychiatric traits. Summary statistics related to anthropologic characteristics included 28 personality, 16 lifestyle/events, 7 diet, 7 physical, 6 sleep, 4 neuroimaging, and 3 longevity traits.

Following exclusion of phecodes with insufficient data, we retained 1,405 phenotypes for the PheWAS analysis. Disease phenotypes included 149 circulatory system, 131 digestive, 129 genitourinary, 123 endocrine/metabolic, 120 neoplasms, 96 musculoskeletal, 92 sense organs, 77 injuries & poisonings, 77 respiratory, 72 dermatological, 71 neurological, 70 psychiatric disorders, 47 infectious diseases, 45 hematopoietic, 36 symptoms, 35 congenital anomalies, and 35 pregnancy complications.

We explored associations with cannabis PGS using 314 LabWAS phenotypes. LabWAS phenotypes included 74 immune, 68 metabolic, 56 blood, 24 cardiovascular, 23 urinary, 20 endocrine, 16 toxicology/pharmacology, 14 other, 6 cancer, 6 kidney, 6 liver, and 1 OB/GYN.

**Polygenic Score Prediction in *All of Us***

The following concept ID codes available in *All of Us* (AoU) were used to create cohorts of participants with cannabis use traits of interest:

| Concept ID(s) | Question/Criteria |
|---|---|
| *Lifetime Cannabis Use* | |
| 1585636 | Answered MARIJUANA to the question “In your LIFETIME, which of the following substances have you ever used?” |
| *Daily Cannabis Use* | |
| 1585650 | Answered DAILY to the question "In the PAST THREE MONTHS, how often have you used Marijuana (cannabis, pot, grass, hash, weed, etc.)?" |
| *Problematic Cannabis Use* | |
| 434327 | Cannabis abuse EHR |
| 440387 | Cannabis dependence EHR |
| 440996 | Cannabis dependence in remission EHR |
| 433452 | Cannabis dependence, continuous EHR |
| 437838 | Cannabis dependence, episodic EHR |
| 4323639 | Cannabis misuse EHR |
| 4103419 | Nondependent cannabis abuse EHR |
| 435231 | Nondependent cannabis abuse in remission EHR |
| 434019 | Nondependent cannabis abuse, continuous EHR |
| 434328 | Nondependent cannabis abuse, episodic EHR |

To calculate liability scale $R^2$ estimates for polygenic prediction based on Lee et al. [14], the estimated prevalence of cannabis use traits in the US population was obtained from the literature for individuals over 18 years old.

| Trait | Estimate (%) | Criteria |
|---|---|---|
| Lifetime cannabis use[15] | 50.30 | Answering yes to the question "Have you ever, even once, used marijuana or any cannabis product?" in the National Survey on Drug Use and Health Survey |
| Daily or almost daily cannabis use[15] | 25.10 | Reporting using marijuana on ≥20 days in the past month in the National Survey on Drug Use and Health Survey |
| DSM-5 CUD[16] | 6.27 | ≥2 out of 11 diagnostic criteria within a 12-month period in the, aggregated with prior diagnosis history in the National Epidemiologic Survey on Alcohol and Related Conditions-III |

### Supplementary Figures

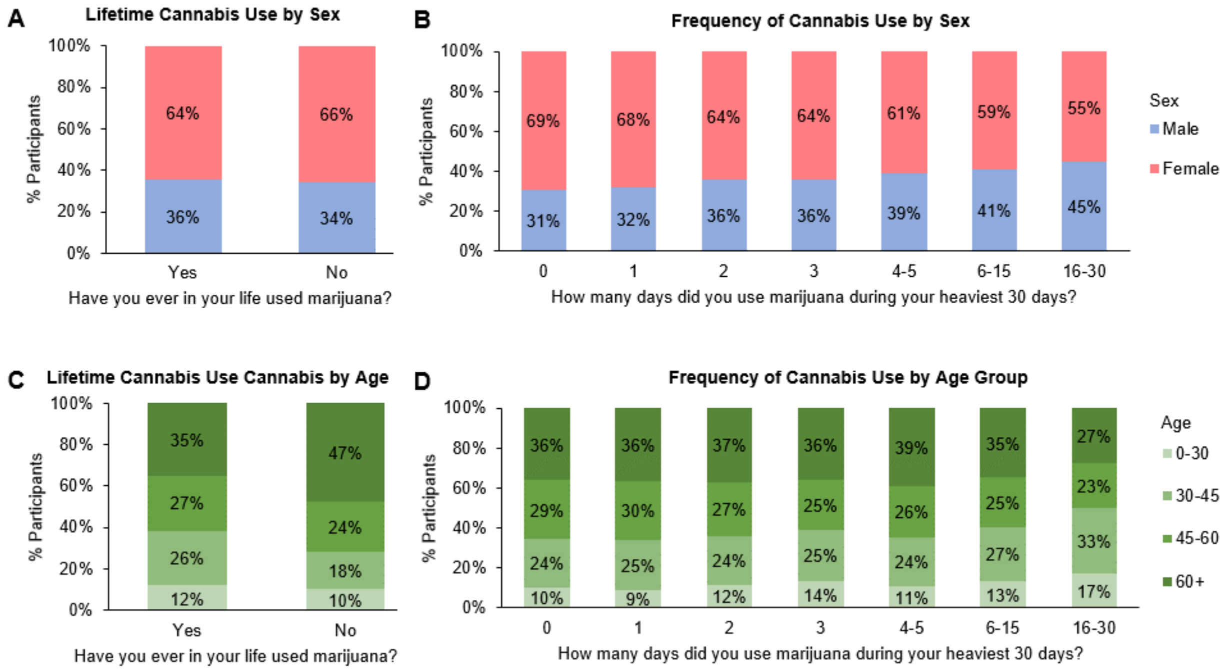


**Supplementary Figure 1.** Participant cannabis use by sex and age group. **A)** Proportion of reported lifetime cannabis use by sex. **B)** Proportion of frequency of cannabis use by sex. **C)** Proportion of reported lifetime cannabis use by age groups. **D)** Proportion of reported frequency of cannabis use by age group.

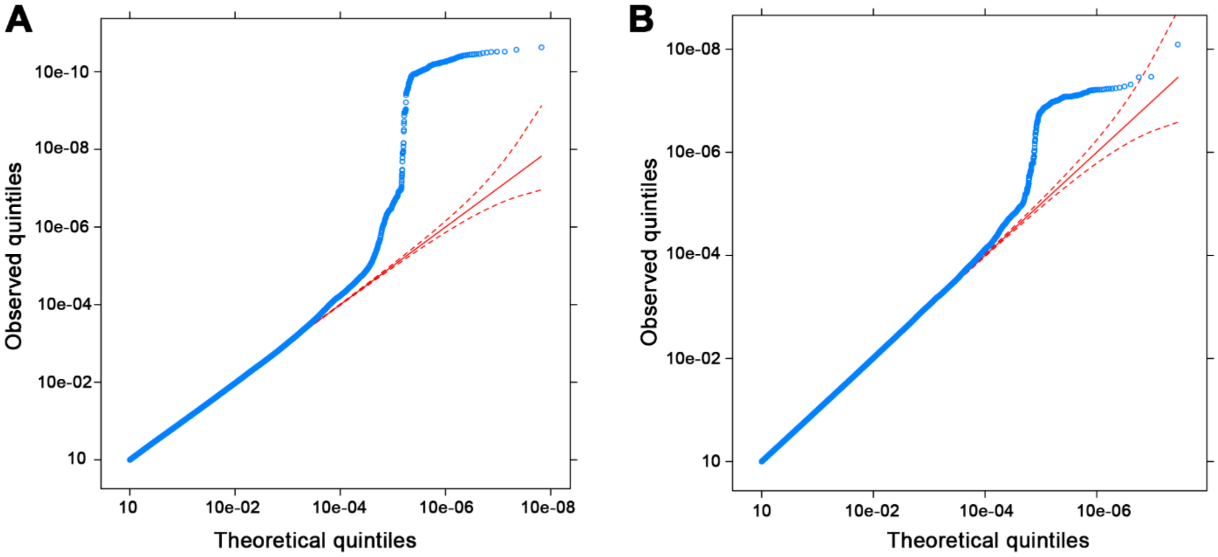


**Supplementary Figure 2.** Q-Q plots for **A)** lifetime cannabis use and **B)** frequency of cannabis use.

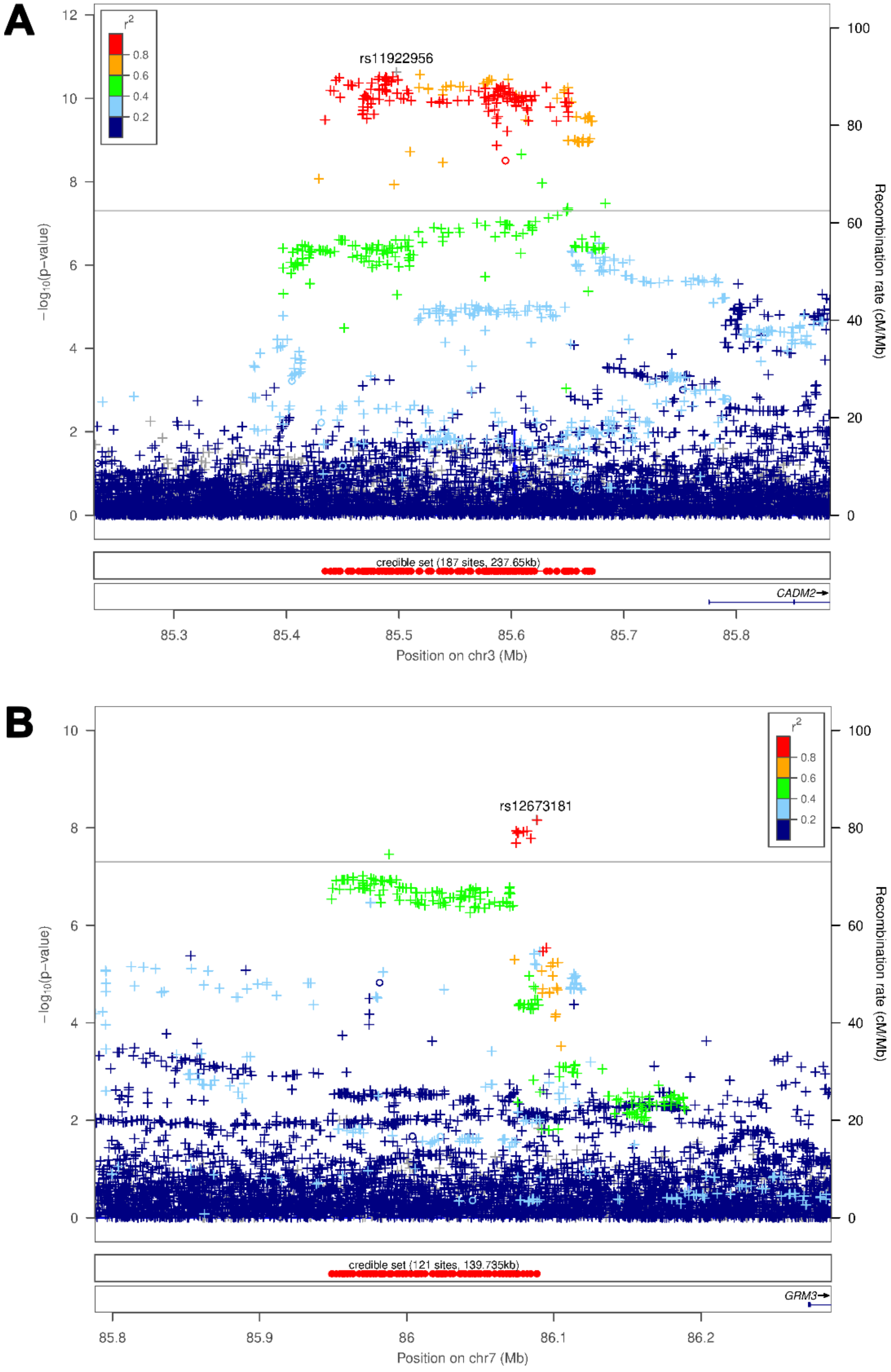
A
B
rs11922956
rs12673181
credible set (187 sites, 237.65kb)
credible set (121 sites, 139.735kb)
CADM2
GRM3
Position on chr3 (Mb)
Position on chr7 (Mb)
−log10(p−value)
Recombination rate (cM/Mb)

142

**Supplementary Figure 3.** Locus zoom plot focusing on SNP **A)** rs11922956 on chromosome 3 and **B)** rs12673181 on chromosome 7. These plots were generated using LocusZoom [17]. The -$\log_{10}$(*p*-value) is shown on the left *y*-axis; position in Mb is on the *x*-axis. Recombination rates (expressed in centiMorgans cM per Mb; NCBI Build GRCh37; highlighted in blue) are shown on the right *y*-axis. Pairwise linkage disequilibrium ($r^2$) of each SNP with the top SNP in the region is indicated by its color. Crossed points represent imputed SNPs, circles represent directly genotyped SNPs.

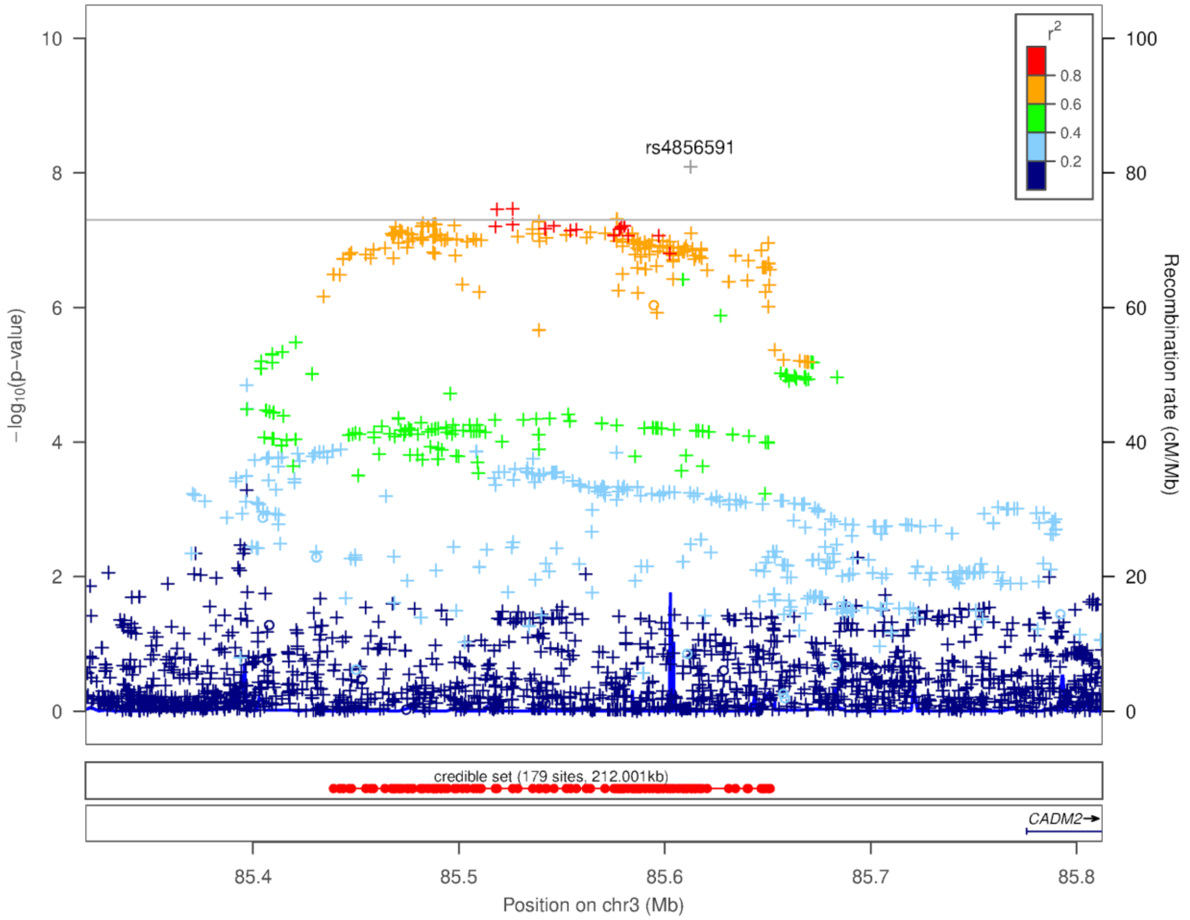


**Supplementary Figure 4.** Locus zoom plot focusing on SNP rs4856591 on chromosome 3. This plot was generated using LocusZoom [17]. The -$\log_{10}$($p$-value) is shown on the left $y$-axis; position in Mb is on the $x$-axis. Recombination rates (expressed in centiMorgans cM per Mb; NCBI Build GRCh37; highlighted in blue) are shown on the right $y$-axis. Pairwise linkage disequilibrium ($r^2$) of each SNP with the top SNP in the region is indicated by its color. Crossed points represent imputed SNPs, circles represent directly genotyped SNPs.

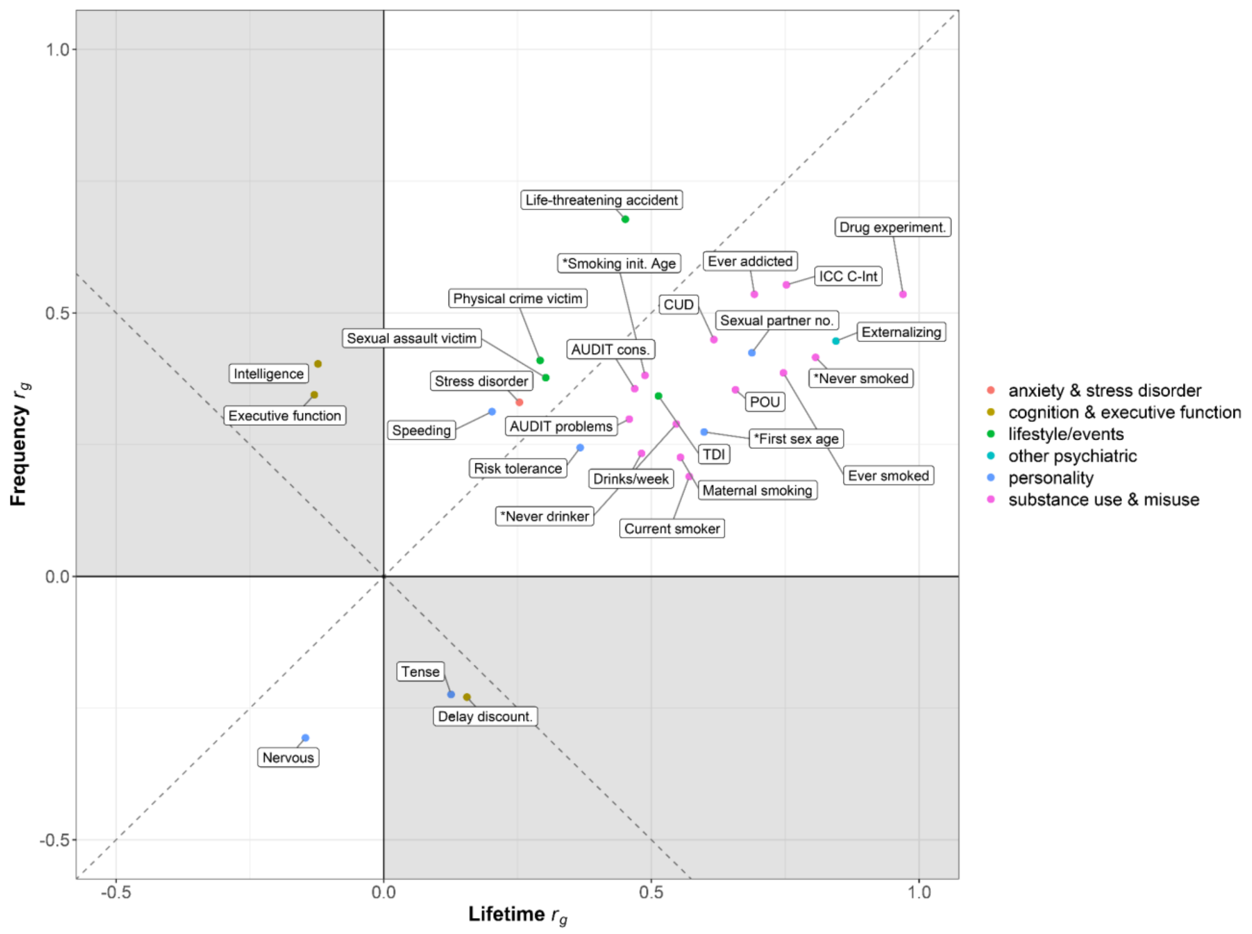


**Supplementary Figure 5.** Comparison of shared FDR-significant genetic correlations between lifetime cannabis use and frequency of cannabis use. Traits with discordant genetic correlation directions of effect located in gray quadrants. Dotted lines represent point of full linearity between lifetime and frequency of cannabis use. *reverse coded traits.

### References


1. Bryc, K., Durand, E.Y., Macpherson, J.M., Reich, D., and Mountain, J.L. (2015). The genetic ancestry of African Americans, Latinos, and European Americans across the United States. Am J Hum Genet *96*, 37-53. 10.1016/j.ajhg.2014.11.010.
2. National Academies of Sciences, Engineering, and Medicine Health and Medicine Division, Division of Behavioral and Social Sciences and Education, Board on Health Sciences Policy, Committee on Population, Committee on the Use of Race, and Ethnicity, and Ancestry as Population Descriptors in Genomics Research (2023). In Using Population Descriptors in Genetics and Genomics Research: A New Framework for an Evolving Field. 10.17226/26902.
3. Durand, E.Y., Do, C.B., Mountain, J.L., and Macpherson, J.M. (2014). Ancestry Composition: A Novel, Efficient Pipeline for Ancestry Deconvolution. bioRxiv. [Preprint]. 10.1101/010512.
4. Eriksson, N., Macpherson, J.M., Tung, J.Y., Hon, L.S., Naughton, B., Saxonov, S., Avey, L., Wojcicki, A., Pe'er, I., and Mountain, J. (2010). Web-based, participant-driven studies yield novel genetic associations for common traits. PLoS Genet *6*, e1000993. 10.1371/journal.pgen.1000993.
5. Hyde, C.L., Nagle, M.W., Tian, C., Chen, X., Paciga, S.A., Wendland, J.R., Tung, J.Y., Hinds, D.A., Perlis, R.H., and Winslow, A.R. (2016). Identification of 15 genetic loci associated with risk of major depression in individuals of European descent. Nat Genet *48*, 1031-1036. 10.1038/ng.3623.
6. Barkley-Levenson, A.M., Lagarda, F.A., and Palmer, A.A. (2018). Glyoxalase 1 (GLO1) Inhibition or Genetic Overexpression Does Not Alter Ethanol's Locomotor Effects: Implications for GLO1 as a Therapeutic Target in Alcohol Use Disorders. Alcohol Clin Exp Res *42*, 869-878. 10.1111/acer.13623.

7. Distler, M.G., Plant, L.D., Sokoloff, G., Hawk, A.J., Aneas, I., Wuenschell, G.E., Termini, J., Meredith, S.C., Nobrega, M.A., and Palmer, A.A. (2012). Glyoxalase 1 increases anxiety by reducing GABAA receptor agonist methylglyoxal. J Clin Invest *122*, 2306-2315. 10.1172/JCI61319.

8. Henn, B.M., Hon, L., Macpherson, J.M., Eriksson, N., Saxonov, S., Pe'er, I., and Mountain, J.L. (2012). Cryptic distant relatives are common in both isolated and cosmopolitan genetic samples. PLoS One *7*, e34267. 10.1371/journal.pone.0034267.

9. Sanchez-Roige, S., Fontanillas, P., Elson, S.L., 23andMe Research Team, Pandit, A., Schmidt, E.M., Foerster, J.R., Abecasis, G.R., Gray, J.C., de Wit, H., et al. (2018). Genome-wide association study of delay discounting in 23,217 adult research participants of European ancestry. Nat Neurosci *21*, 16-18. 10.1038/s41593-017-0032-x.

10. Sanchez-Roige, S., Fontanillas, P., Jennings, M.V., Bianchi, S.B., Huang, Y., Hatoum, A.S., Sealock, J., Davis, L.K., Elson, S.L., 23andMe Research Team, and Palmer, A.A. (2021). Genome-wide association study of problematic opioid prescription use in 132,113 23andMe research participants of European ancestry. Mol Psychiatry *26*, 6209-6217. 10.1038/s41380-021-01335-3.

11. Sanchez-Roige, S., Jennings, M.V., Thorpe, H.H.A., Mallari, J.E., van der Werf, L.C., Bianchi, S.B., Huang, Y., Lee, C., Mallard, T.T., Barnes, S.A., et al. (2023). CADM2 is implicated in impulsive personality and numerous other traits by genome- and phenome-wide association studies in humans and mice. Transl Psychiatry *13*, 167. 10.1038/s41398-023-02453-y.

12. Sanchez-Roige, S., Palmer, A.A., Fontanillas, P., Elson, S.L., 23andMe Research Team, Substance Use Disorder Working Group of the Psychiatric Genomics Consortium, Adams, M.J., Howard, D.M., Edenberg, H.J., Davies, G., et al. (2019). Genome-Wide Association Study Meta-Analysis of the Alcohol Use Disorders Identification Test

(AUDIT) in Two Population-Based Cohorts. Am J Psychiatry *176*, 107-118. 10.1176/appi.ajp.2018.18040369.

13. Fuchsberger, C., Abecasis, G.R., and Hinds, D.A. (2015). minimac2: faster genotype imputation. Bioinformatics *31*, 782-784. 10.1093/bioinformatics/btu704.

14. Lee, S.H., Goddard, M.E., Wray, N.R., and Visscher, P.M. (2012). A better coefficient of determination for genetic profile analysis. Genet Epidemiol *36*, 214-224. 10.1002/gepi.21614.

15. Substance Abuse and Mental Health Services Administration. (2023). Results from the 2022 National Survey on Drug Use and Health: Detailed tables.

16. Hasin, D.S., Kerridge, B.T., Saha, T.D., Huang, B., Pickering, R., Smith, S.M., Jung, J., Zhang, H., and Grant, B.F. (2016). Prevalence and Correlates of DSM-5 Cannabis Use Disorder, 2012-2013: Findings from the National Epidemiologic Survey on Alcohol and Related Conditions-III. Am J Psychiatry *173*, 588-599. 10.1176/appi.ajp.2015.15070907.

17. Pruim, R.J., Welch, R.P., Sanna, S., Teslovich, T.M., Chines, P.S., Gliedt, T.P., Boehnke, M., Abecasis, G.R., and Willer, C.J. (2010). LocusZoom: regional visualization of genome-wide association scan results. Bioinformatics *26*, 2336-2337. 10.1093/bioinformatics/btq419.

18. Pasman, J.A., Verweij, K.J.H., Gerring, Z., Stringer, S., Sanchez-Roige, S., Treur, J.L., Abdellaoui, A., Nivard, M.G., Baselmans, B.M.L., Ong, J.S., et al. (2018). GWAS of lifetime cannabis use reveals new risk loci, genetic overlap with psychiatric traits, and a causal influence of schizophrenia. Nat Neurosci *21*, 1161-1170. 10.1038/s41593-018-0206-1.

19. Levey, D.F., Galimberti, M., Deak, J.D., Wendt, F.R., Bhattacharya, A., Koller, D., Harrington, K.M., Quaden, R., Johnson, E.C., Gupta, P., et al. (2023). Multi-ancestry genome-wide association study of cannabis use disorder yields insight into disease

biology and public health implications. Nat Genet *55*, 2094-2103. 10.1038/s41588-023-01563-z.
